# Trisomy 21 Drives *ADARB1* Overexpression and Premature RNA Recoding in the Developing Fetal Brain

**DOI:** 10.1101/2025.11.13.25339504

**Authors:** Michael S. Breen, Andy Yang, Xuran Wang, Miguel Rodriguez de los Santos, Ran Tao, Daniel R. Weinberger, Joel E. Kleinman, Kalina Mihova, Gergana Stancheva, Sylvia Savova, Radka Kaneva, Violeta Dimitrova, Vladimir Vladimirov, Thomas M. Hyde, Joseph D. Buxbaum

**Author notes:** Shared first authorship (Michael S. Breen, Andy Yang, Xuran Wang).

## Abstract

Understanding how chromosome 21 gene dosage contributes to neurodevelopmental and systemic phenotypes in trisomy 21 (T21) remains a fundamental challenge. We performed transcriptome-wide RNA sequencing on fetal cortical and hippocampal tissues from 20 T21 cases and 27 euploid controls collected between 13–22 weeks post-conception, a critical period for human brain development. Differential expression analysis revealed 572 dysregulated genes in the prefrontal cortex and 519 in the hippocampus (FDR < 5%), with significant enrichment for chromosome 21 genes. Functional enrichment analyses highlighted disruptions in neurodevelopmental, synaptic, and immune-related pathways. Among the most strongly dysregulated genes was *ADARB1*, a chromosome 21-encoded RNA editing enzyme, whose overexpression in T21 fetal brain was associated with increased adenosine-to-inosine (A-to-I) editing, including recoding sites in *GRIA2* (p.R764G), *GRIA3* (p.R775G), and *GRIK2* (p.Y571C, p.Q621R). A meta-analysis incorporating nine independent transcriptomic datasets spanning early embryonic and progenitor cell types validated robust chromosome 21 dosage effects, including consistent *ADARB1* overexpression. Extending these findings, a meta-analysis of A-to-I editing across datasets revealed widespread over-editing at 3′UTRs and at *GRIA3* (p.R775G), a site critical for AMPA receptor desensitization. Together, these results implicate dysregulated RNA editing driven by *ADARB1* overexpression as a post-transcriptional mechanism contributing to fetal neuropathology in T21 and provide a framework for understanding the broader molecular consequences of chromosome 21 dosage sensitivity during brain development.

## INTRODUCTION

Trisomy 21 (T21), the genetic hallmark of Down syndrome (DS), results in the triplication and overexpression of genes on chromosome 21, disrupting key neurodevelopmental processes^1^. Although the genetic cause of DS is well defined, the molecular mechanisms by which T21 disrupts fetal brain development remain incompletely understood and mechanistically unresolved. Transcriptomic analyses across developmental stages and brain regions have revealed widespread dysregulation of gene expression, implicating both chromosome 21 genes and broader genome-wide effects^2–4^. In fetal brain from individuals with DS, studies have identified spatially and temporally specific transcriptional changes, including impaired oligodendrocyte differentiation and disrupted myelination^3^, suggesting a mechanistic link to abnormalities in white matter integrity and neural circuit formation. In postnatal and adult DS brains, transcriptional dysregulation persists, extending to pathways involved in oxidative stress responses, cytoskeletal organization, and immune signaling^5–7^.

Emerging evidence suggests that post-transcriptional regulation, particularly RNA editing, may play a critical role in the neurodevelopmental phenotypes of DS. *ADARB1*, a chromosome 21-encoded enzyme that catalyzes adenosine-to-inosine (A-to-I) editing, modulates RNA sequence diversity, neuronal excitability, and synaptic function during brain development^8,9^. A dosage-sensitive gene, *ADARB1* is consistently overexpressed in DS cellular models and *in vivo* tissues^4,10–13^. However, the extent to which this alters RNA editing activity in the developing human brain remains unclear. By contrast, precise editing disruptions at recoding sites have been implicated in related neurodevelopmental and neuropsychiatric disorders, including Alzheimer’s disease¹ , autism spectrum disorder¹ , and schizophrenia¹ , where they impair synaptic signaling and circuit maturation. Whether similar editing-dependent mechanisms contribute to neurodevelopmental abnormalities in DS remains an open question.

In parallel with transcriptional and post-transcriptional disruptions, immune dysregulation has emerged as a central and persistent feature of DS. The triplication of interferon receptor genes (*IFNAR1*, *IFNAR2*, and *IFNGR2*) on chromosome 21 amplifies interferon signaling pathways, leading to chronic immune activation and sustained upregulation of interferon-stimulated genes^17–20^. While *ADAR*, particularly the interferon-inducible p150 isoform, is canonically upregulated by inflammatory signals^21,22^, our prior transcriptomic analyses of non-DS human brain tissues revealed that interferon activation is also associated with increased *ADARB1* expression^23^, suggesting that *ADARB1* may also be responsive to immune tone *in vivo*. This is particularly relevant in DS, where gene dosage imbalance uniquely co-occurs with heightened interferon activity, creating a biologically distinct environment that may alter RNA editing programs. The intersection of interferon signaling, gene triplication, and post-transcriptional control represents a largely unexplored axis of pathology with the potential to disrupt neurodevelopmental trajectories and impair synaptic function.

Despite these insights, key gaps remain in our understanding of DS molecular pathology during early brain development. To date, only two transcriptomic studies have examined DS fetal brain tissue (using microarrays): one analyzing 15 DS and 15 control brains^3^, and another profiling 9 DS and 8 control brains^4^. While foundational, these studies, along with independent hiPSC-derived neuron models^10–13^, have been limited in both scale and resolution and were not designed to assess post-transcriptional regulation such as RNA editing. Additional efforts have focused primarily on postnatal or adult tissues using targeted PCR or, more recently, single-nucleus RNA sequencing (snRNA-seq) ^3,4,7,24^. Although snRNA-seq provides cell type-specific resolution, it is not well suited for genome-wide, quantitative analysis of editing levels in fetal brain. Furthermore, no study has systematically examined how chromosome 21 gene dosage, interferon signaling, and RNA editing intersect during early brain development. Addressing these gaps requires high-resolution transcriptomic profiling across key developmental windows and brain regions.

Here, we address these critical gaps by performing transcriptome-wide RNA sequencing of fetal prefrontal cortical and hippocampal tissues from 20 individuals with T21 and 27 euploid controls, spanning 13-22 weeks post-conception, a key window for human brain maturation (**Figure 1**). By meta-analyzing these data with 9 independent transcriptomic datasets, we define robust and reproducible chromosome 21 dosage effects and reveal convergent transcriptional and post-transcriptional disruptions across diverse developmental stages and cellular contexts. Focusing on RNA editing, we demonstrate that *ADARB1* is a dosage-sensitive regulator of A-to-I editing in the developing brain, driving premature and excessive recoding at key genes during neurodevelopment. Together, these findings establish RNA editing dysregulation as a novel and conserved mechanism of T21 neuropathology and provide a framework for dissecting post-transcriptional contributions to altered brain development in DS.

**Figure 1.**
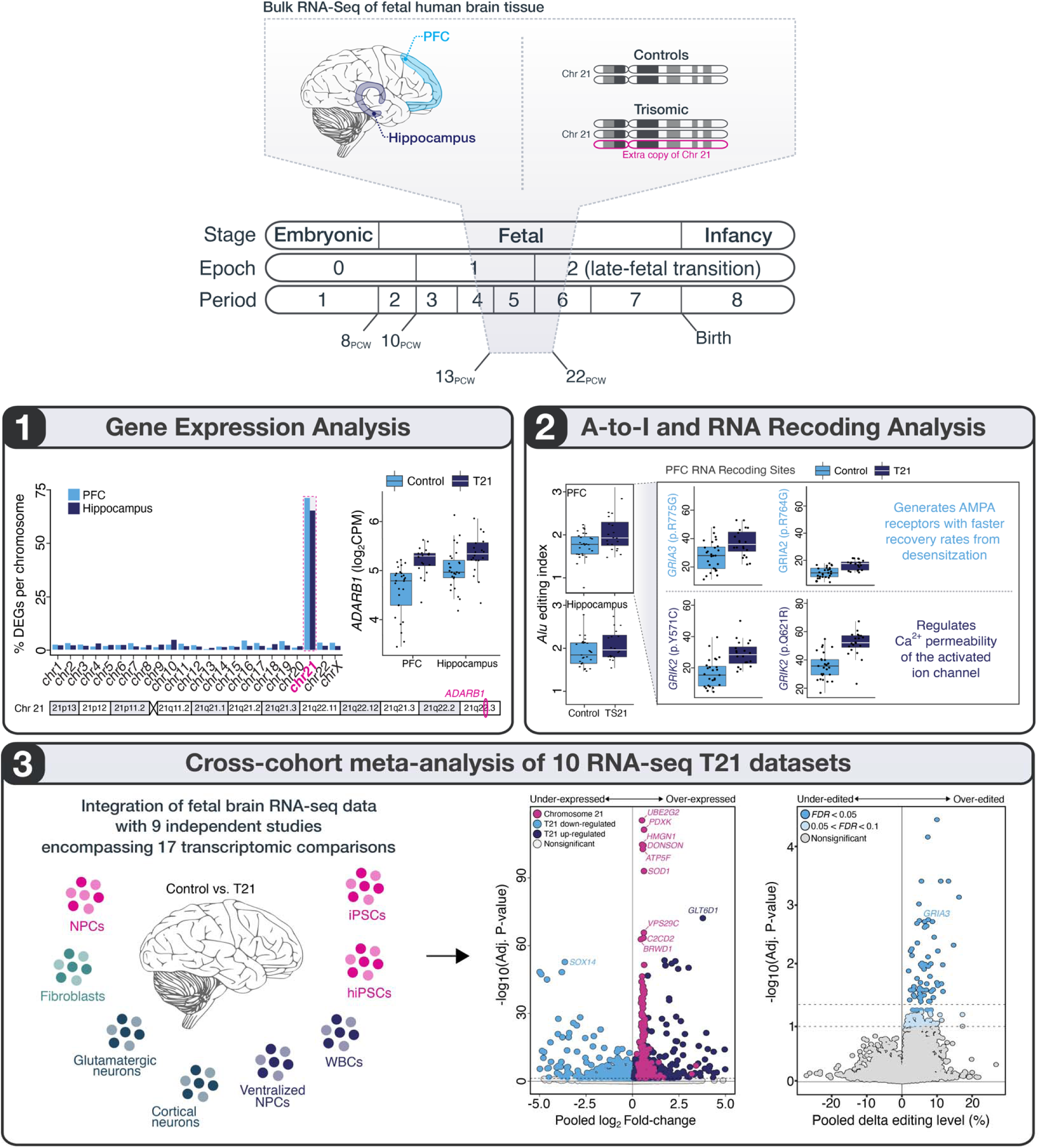
Overview of study design and analytical framework. We performed deep bulk RNA sequencing of paired prefrontal cortex (PFC) and hippocampal tissue from 20 mid-gestation fetuses with trisomy 21 (T21) and 21 age- and sex-matched euploid controls (13–22 post-conception weeks [PCW]). A multi-layered analytical pipeline was applied to: (1) Quantify differential gene expression and assess selective gene dosage effects on chromosome 21; (2) profile global and site-specific adenosine-to-inosine (A-to-I) editing, including recoding events that alter protein sequence; and (3) Perform a harmonized meta-analysis of ten RNA-seq datasets to identify reproducibly dysregulated genes and editing sites in T21.

## RESULTS

We generated deep bulk RNA-sequencing profiles (34.5M ± 5.4M reads/sample) from fetal prefrontal cortex (PFC) and hippocampal tissues of individuals with Trisomy 21 (T21; n = 20) and age-, sex- and ancestry-matched euploid controls (n = 27) (**Table S1**). To assess global transcriptomic variation, we initially applied a unified normalization strategy across all samples, quantifying expression for 16,737 genes. Rigorous quality control analyses were then conducted to identify key sources of variability in the dataset (**Figures S1–S3**). First, cell type deconvolution revealed similar estimated neuronal proportions in PFC (∼62.2%) and hippocampus (∼62.1%), with no significant differences between T21 and control groups (**Figure S1**). Next, principal component analysis (PCA) highlighted neuronal content as the dominant axis of variation (PC1; 29.2% variance explained), while PC2 (17.8% variance) clearly separated PFC and hippocampal tissues (**Figure S2**). To formally assess regional differences, we performed differential expression analysis between brain regions and identified 9,163 differentially expressed genes at FDR < 5%, accounting for ∼54% of the transcriptome (**Figure S3**). Based on these marked region-specific signatures, we conducted all downstream analyses independently for PFC and hippocampus using tailored, region-specific modeling strategies (*see Methods*).

### Transcriptomic effects of trisomy 21 during early fetal brain development

To identify critical covariates for inclusion in our comparative analysis of T21 individuals and controls within each region, we used a linear mixed model to quantify the fraction of expression variance attributable to known clinical and technical factors, including neuronal cell type proportion, percentage of mRNA reads (defined as reads mapping to exons and untranslated regions), gestational age, RNA integrity number (RIN), T21 status, and sex, for each gene, separately within PFC and hippocampus (**Figure S4**). Collectively, these variables accounted for ∼57.1% of transcriptome-wide variation in the PFC and ∼34.9% in the hippocampus. In the PFC, estimated neuronal cell type proportions exerted the largest effect, explaining a median 25.1% of the observed variation, followed by the percentage of mRNA reads (6.2%), gestational age (2.4%), and RIN (0.4%), with remaining factors contributing modestly. Expression variation attributable to T21 itself was detectable, particularly in a subset of genes located on chromosome 21, confirming the robustness of our approach. Similar patterns of variance decomposition were observed independently in the hippocampus (**Figure S4E–H**).

We next performed differential expression analysis comparing individuals with T21 and controls, adjusting for the possible influence of estimated neuronal cell type proportions, RIN, gestational age, sex, and percentage of mRNA reads, within each region. This analysis identified 572 differentially expressed genes (DEGs) in the PFC and 519 in the hippocampus at transcriptome-wide significance (FDR < 5%) (**Figure 2A–B, Figure S5**, **Table S2**). While gene expression changes in T21 were positively correlated across regions (*r*=0.40), the DEG profiles were largely region specific. Only 155 genes were consistently dysregulated in both the PFC and hippocampus, most of which were located on chromosome 21 (**Figure 2C-D**). Among the DEGs, there was significant enrichment of upregulated chromosome 21 genes in individuals with T21 in both the PFC (n_genes_=122, *p*=6.7×10^-142^, one-sided FET) and hippocampus (n_genes_=118, *p* =8.6×10^-137^, one-sided FET) (**Figure 2C,E**), reflecting the canonical gene dosage effects associated with T21.

**Figure 2.**
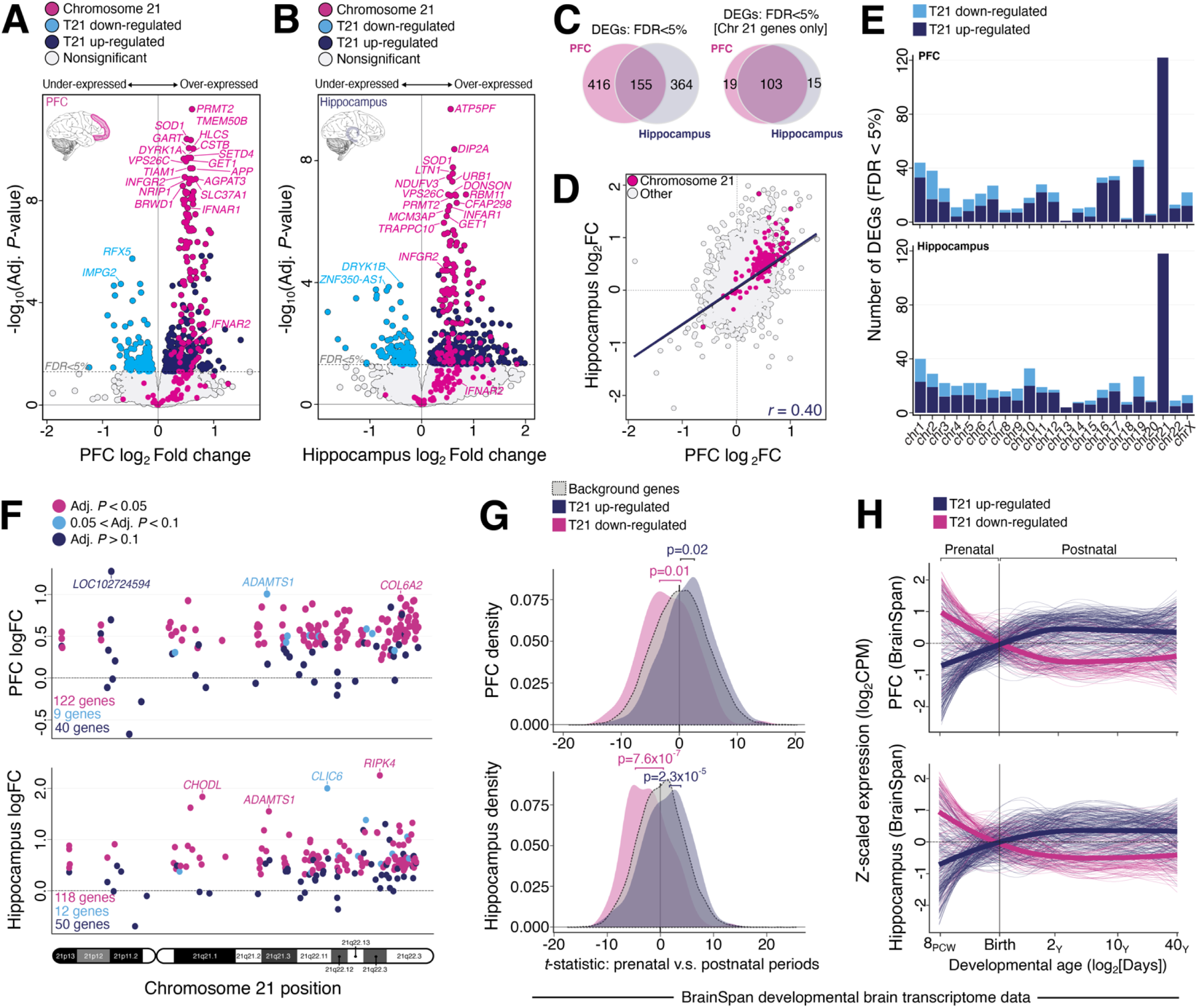
Transcriptome-wide differential expression and developmental expression bias in T21 fetal brain. Differential expression analyses comparing T21 and control samples are shown for the prefrontal cortex (PFC; **A**) and hippocampus (**B**), with each gene plotted by log fold-change (x-axis) and –log adjusted p-value (y-axis); points are colored to indicate upregulated, downregulated, and chromosome 21–encoded genes. (**C**) Venn diagrams display the overlap of all differentially expressed genes (DEGs) (left) and DEGs located on chromosome 21 (right) between PFC and hippocampus. (**D**) Genome-wide concordance of expression changes between regions is shown by plotting log fold-changes in PFC (x-axis) against hippocampus (y-axis); Spearman correlation coefficient (ρ) quantifies regional concordance. (**E**) Barplots show the number of DEGs (FDR < 5%) per chromosome in PFC (top) and hippocampus (bottom), separated by direction of change. (**F**) Dot plots display expression changes for all genes on chromosome 21, arranged by genomic position (x-axis) for PFC (top) and hippocampus (bottom); colors reflect FDR significance thresholds (FDR < 5%, pink; 5% < FDR < 10%, light blue; FDR > 10%, dark blue). (**G**) A t-statistic was computed summarizing prenatal vs. postnatal expression bias using BrainSpan data from matching anatomical regions (PFC and hippocampus). Next, we quantified the distribution of the t-statistics among T21-upregulated and downregulated genes in the PFC (top) and hippocampus (bottom), revealing a prenatal bias among under-expressed genes (p = 0.02 and p = 2.3 × 10 ) and a postnatal bias among over-expressed genes (p = 0.01 and p = 7.6 × 10 ). (**H**) Normalized expression trajectories of T21-upregulated and downregulated genes were plotted across ten developmental stages in the BrainSpan bulk RNA-seq using matching PFC (top) and hippocampus (bottom) tissues, illustrating typical temporal shifts in gene expression trajectories.

Despite the strong enrichment for upregulated chromosome 21 genes, we did not observe a universal dosage effect across the entire chromosome. In the PFC, 40 genes on chromosome 21 showed no significant expression changes between T21 cases and controls, with a similar pattern observed for 50 genes in the hippocampus (**Figure 2F**). These genes were distributed relatively evenly across the chromosome. Notably, chromosome 21 genes that were significantly upregulated in T21 cases were more highly expressed in T21 and control samples compared to non-significant genes (PFC: fold-change = 2.29, *p* = 1.1×10 ¹ ; hippocampus: fold-change = 2.31, *p* = 1.3×10 ¹¹; Wilcoxon rank-sum test) (**Figure S6**). This suggests that gene dosage sensitivity in the T21 brain preferentially affects genes that are already more actively transcribed, while lower-expressed genes may evade dysregulation, potentially due to tighter transcriptional control. Among the dysregulated chromosome 21 genes, we highlight increased expression of key interferon signaling components, including interferon gamma receptor 2 (*IFNGR2*) and interferon alpha/beta receptor subunits 1 and 2 (*IFNAR1*, *IFNAR2*), in both the PFC (FDR *p* = 6.7 × 10 ) and hippocampus (FDR *p* = 3.7 × 10 ²). These genes, which canonically mediate interferon signaling in immune contexts, have also been implicated in brain development and glial function, raising the possibility that their overexpression contributes to immune-related transcriptional changes in the T21 brain. Additionally, we observed upregulation of several collagen genes critical for brain extracellular matrix integrity (*COL6A1*, *COL6A2*, *COL18A1*) in both regions (PFC FDR *p* = 1.4 × 10 ²; hippocampus FDR *p* = 2.5 × 10 ²), as well as increased expression of genes involved in ligase activity (*ATP5PF*, *ATP5PO*, *GART*, *HLCS*, *DIP2A*) (PFC FDR *p* = 2.0 × 10 ²; hippocampus FDR *p* = 3.7 × 10 ²).

### Developmental imbalance of fetal gene expression patterns in individuals with T21

A substantial proportion of dysregulated T21 genes reside outside chromosome 21 and likely represent secondary, downstream effects. To assess whether non-chromosome 21 DEGs reflect broader disruptions in developmental timing, we leveraged transcriptomic data from the BrainSpan atlas to define normative gene expression trajectories in the PFC and hippocampus across prenatal and postnatal stages (**Table S3**). For each gene, we computed a *t*-statistic capturing its expression bias toward prenatal or postnatal timepoints in typical development (*see Methods*). We then tested whether T21-overexpressed and T21-underexpressed genes were disproportionately drawn from genes with strong developmental timing biases, by comparing their BrainSpan-derived *t*-statistic distributions within each brain region. This analysis revealed a striking asymmetry: genes downregulated in T21 were significantly enriched for prenatally expressed genes (PFC: *p* = 0.02; hippocampus: *p* = 2.3 × 10 ; Mann-Whitney U), whereas upregulated genes were enriched for postnatally expressed genes (PFC: *p* = 0.01; hippocampus: *p* = 7.6 × 10 ; Mann-Whitney U) (**Figure 2G–H**). These findings suggest that T21 disrupts the temporal coordination of gene expression, prematurely activating postnatal programs while attenuating prenatal ones during early brain development.

### Systems-level dysregulation of gene expression in T21

To further characterize the biological processes underlying this temporal dysregulation, we performed functional enrichment analysis of differentially expressed genes located outside chromosome 21 (**Table S4**). Using a competitive gene set ranking framework, we identified significant enrichment of down-regulated genes involved in mitochondrial translation and mRNA binding in both the PFC and hippocampus (**Figure 3A-B)**, pointing to impaired metabolic function and post-transcriptional regulation during early neurodevelopment. This was accompanied by reduced expression of genes critical for translational initiation and elongation, ribosomal RNA processing, and nonsense-mediated mRNA decay, suggesting broader suppression of protein synthesis and RNA quality control pathways in the T21 fetal brain. In contrast, over-expressed genes were enriched for sodium channel activity across both regions and extracellular matrix (ECM) organization specifically in the hippocampus, including *DCN* (Decorin) and *LTBP4*, which regulate TGF-β signaling in the ECM microenvironment. Together, these results suggest that T21-associated gene dysregulation involves both suppression of early translational and post-transcriptional machinery and premature activation of pathways associated with neuronal excitability and structural maturation.

**Figure 3.**
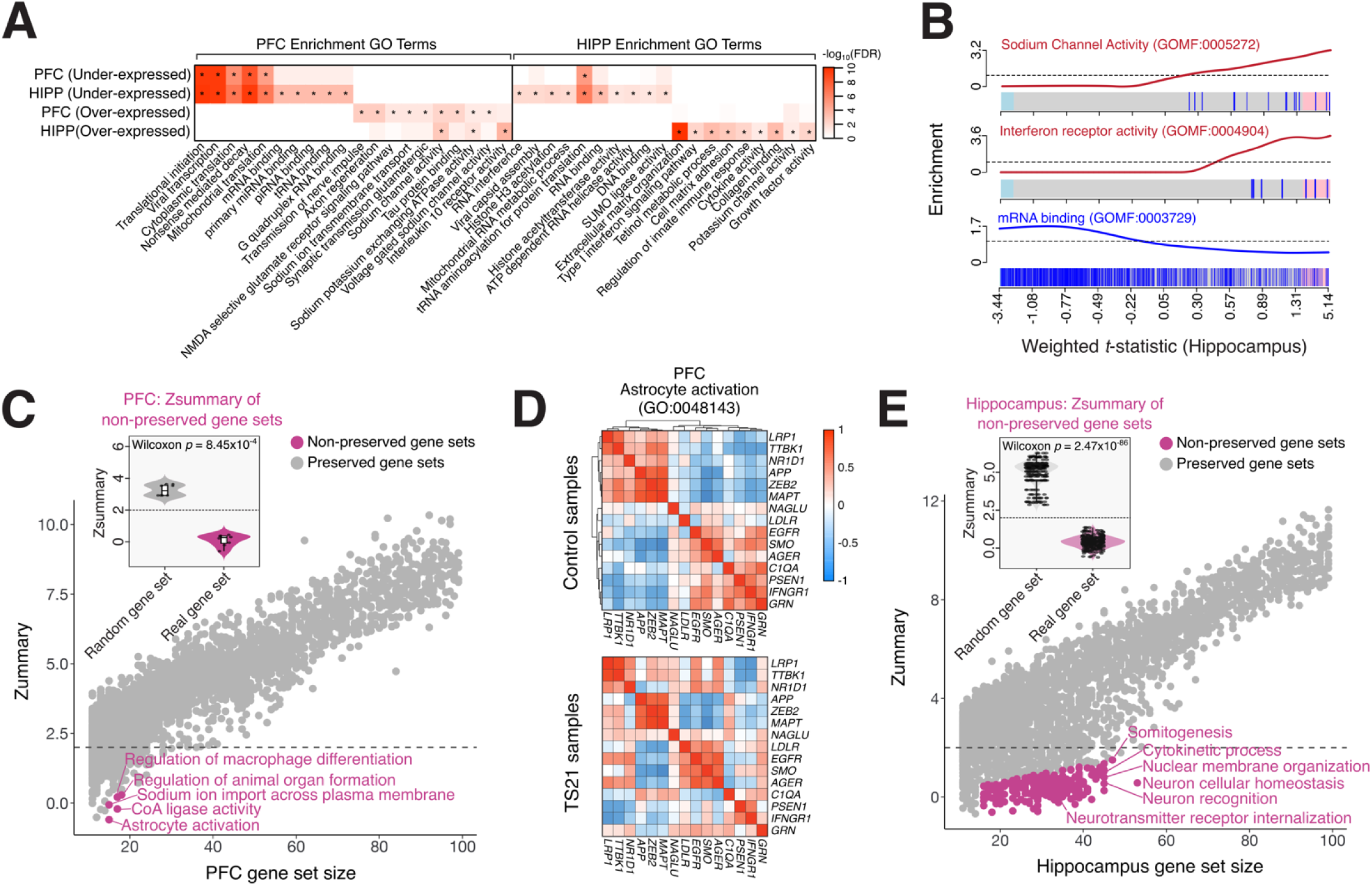
Functional annotation and systems-level disorganization of T21-associated transcriptomic changes. (**A**) Heatmap summarizing Correlation Adjusted MEan RAnk (CAMERA) gene set enrichment analysis, a competitive method that accounts for inter-gene correlation when testing whether predefined gene sets are enriched among differentially expressed genes, based on transcriptome-wide over- and under-expressed genes in T21. The y-axis lists the top five most significantly enriched gene sets (FDR-adjusted p-values) for each category of DEGs within the PFC and hippocampus. (**B**) Enrichment barcode plots for selected gene sets illustrate enrichment patterns across ranked genes (x-axis) sorted by log fold-change in T21. Vertical bars indicate the positions of genes from the target gene set, demonstrating enrichment for sodium channel activity, interferon receptor signaling, and mRNA binding, highlighted examples corresponding to gene sets in panel A. (**C–E**) Gene set preservation analysis assessed whether co-expression patterns of predefined biological gene sets were maintained between T21 and control samples. Preservation was evaluated in the (**C–D**) PFC and (**E**) hippocampus using a combination of Zsummary metrics and empirical permutation testing. Gene sets were classified as non-preserved if they met multiple stringent criteria: low Zsummary1 (<2), lack of statistical significance after FDR correction (p.adj > 0.1), deviation from predicted preservation by ≥4 Z-score units, and significant preservation under random permutation (p.random.adj < 0.05), and are colored coded accordingly. Inset boxplots summarize the distribution of Zsummary scores across flagged non-preserved gene sets. (**D**) As a representative example, correlation matrices of genes within the astrocyte activation gene ontology term are visualized in control samples (top, unsupervised clustering) and T21 samples (bottom, force-ordered to match control structure), illustrating disrupted co-regulatory structure in T21.

While differential expression and functional enrichment analyses identify genes and pathways with altered mean expression levels, they do not capture whether the underlying coordination of gene activity within biological programs is preserved. To assess whether T21 disrupts the structural organization of transcriptional networks, we first applied weighted gene co-expression network analysis (WGCNA) to identify unsupervised gene modules in PFC and hippocampus. This analysis did not reveal major differences in overall module architecture between T21 and control samples (**Figure S7**). We next used a gene set preservation framework to test whether predefined biological gene sets retained coordinated expression across groups, an approach that can detect systems-level dysregulation even in the absence of changes in gene abundance (*see Methods*) (**Table S5**).

In the PFC, eight gene sets exhibited markedly reduced preservation in T21, including those involved in astrocyte activation, sodium ion transport, and macrophage differentiation, suggesting altered regulatory interactions among glial cell types (**Figure 3C-D**). In contrast, the hippocampus showed more widespread co-expression disruption, with 259 gene sets demonstrating poor preservation between individuals with T21 and controls (**Figure 3E-F**). These gene sets clustered into four major domains: (1) neurodevelopment and synaptic plasticity (e.g., *synapse maturation*, *long-term synaptic depression*, *positive regulation of excitatory postsynaptic potential*); (2) chromatin and epigenetic regulation (e.g., *ATP-dependent chromatin remodeler activity*, *chromatin protein adaptor activity*); (3) mitochondrial metabolism (e.g., *NADH dehydrogenase ubiquinone activity*, *oxidoreductase activity*, *NADH metabolic process*); and (4) immune signaling (e.g., *interleukin-4 production*, *regulation of neuroinflammatory response*, *monocyte differentiation*). These findings suggest that T21 disrupts the integrity of gene co-regulation across multiple biological systems, particularly those governing neuronal signaling, energy metabolism, and glial immune responses.

### Meta-analysis across developmental datasets validates chromosome 21 dosage effects in T21

To validate key findings and assess the generalizability of chromosome 21 dosage effects, we conducted a meta-analysis combining our fetal RNA-seq data with nine publicly available RNA-sequencing datasets (**Table 1**). Altogether, we analyzed 108 T21 and 117 euploid control samples from ten datasets, spanning early gestational stages and diverse neural lineages (e.g., iPSCs, NPCs, neurons, fibroblasts), thereby capturing diverse but developmentally relevant cellular states. To minimize inter-study bias, all datasets were reprocessed through a unified RNA-seq analysis pipeline, standardizing quantification and differential expression estimation across studies. For each dataset, a linear model with empirical Bayes moderation was applied to estimate log fold-changes, adjusting for available biological and technical covariates (e.g., age, sex, cell type proportion, sequencing depth). These covariate-adjusted estimates were then synthesized across datasets using a random-effects meta-regression framework to account for between-study heterogeneity. One previously published microarray study of T21 fetal brain^3^ was excluded from our meta-analysis due to technical and analytic limitations. Despite profiling overlapping regions and developmental stages, it showed only modest transcriptome-wide concordance with our RNA-seq data (PFC: *r* = 0.065; hippocampus vs. cerebellum: *r* = 0.11) and weak correlation for chromosome 21 genes (*r* = 0.27 and 0.10, respectively; **Figure S8**, **Table S6**). These discrepancies, along with the platform’s inability to assess RNA editing, precluded its inclusion in our synthesis. While cellular contexts varied across datasets, our goal was to identify genes consistently dysregulated across developmental lineages, rather than those specific to any single cell type. To ensure analytical consistency across platforms and study designs, each dataset was reprocessed from raw FASTQ files using a standardized RNA-seq pipeline for alignment, quantification, and normalization (*see Methods*).

**Table 1.**
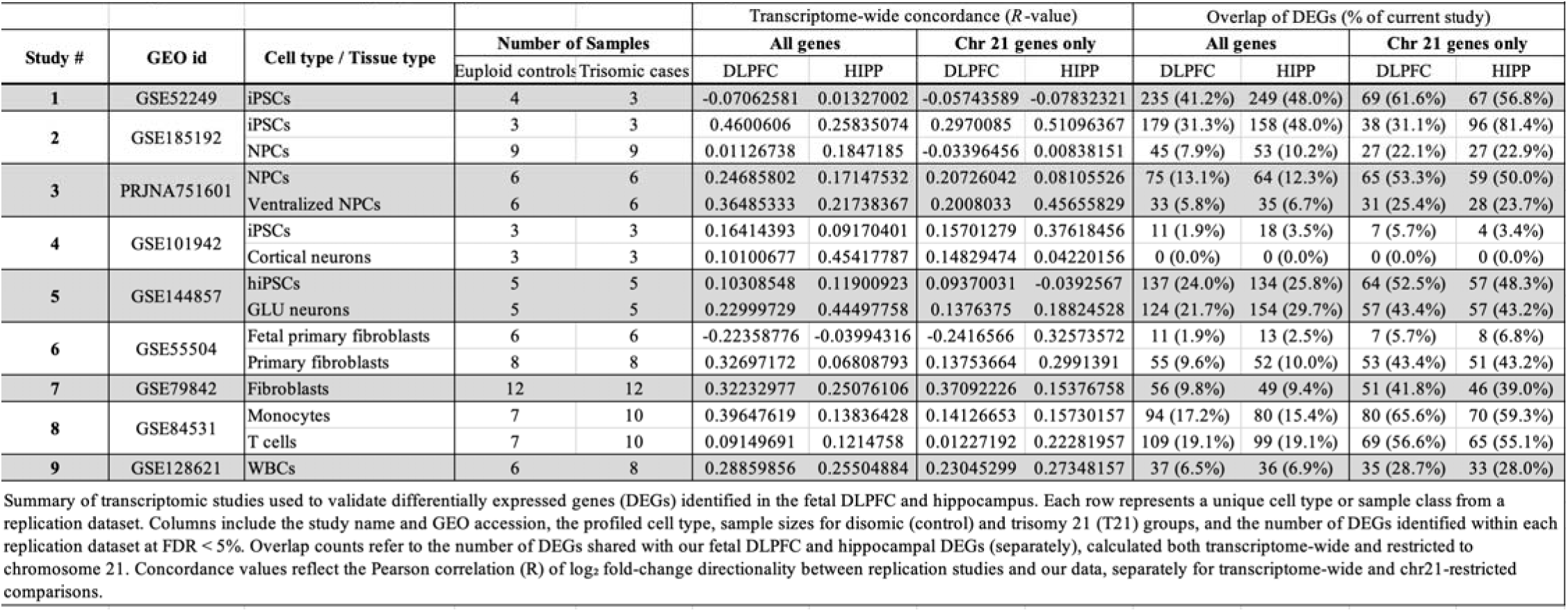
Cross-study validation of differentially expressed genes in Trisomy 21 across developmental and cellular models.

Despite variability in sample sizes and experimental conditions, a consistent pattern emerged: genome-wide fold-change distributions revealed a distinct and reproducible elevation of gene expression on chromosome 21 in all comparisons (**Figure S9**). When plotting log fold changes across the transcriptome, chromosome 21 genes consistently showed an upward shift relative to the rest of the genome, reflecting their dosage-driven overexpression in T21. This global effect, observed across iPSCs, NPCs, neurons, and fibroblasts, reinforces the robustness of chromosome 21 gene dysregulation as a defining feature of T21, regardless of tissue or developmental context. Notably, this effect was not uniformly distributed across the chromosome: only a subset of chromosome 21 genes showed consistent overexpression (**Figure S10**), reinforcing a model of targeted rather than global dosage sensitivity, consistent with emerging concepts of dosage compensation and chromatin architecture.

To formally synthesize findings across datasets, we performed a transcriptome-wide meta-analysis, identifying 543 upregulated and 437 downregulated genes in T21 at strict genome-wide significance (FDR < 1%) (**Figure 4A, Table S7**), reflecting a more conservative threshold appropriate for integrating heterogeneous datasets and reducing false positives. Among the upregulated genes, 149 (27.4%) were located on chromosome 21. The average log fold-change for chromosome 21 genes approximated 0.58 (1.5-fold), consistent with expectations for gene dosage, while variation in effect sizes pointed to selective transcriptional sensitivity across loci (**Figure 4B**). This analysis produced a robust catalogue of consistently dysregulated chromosome 21 genes across diverse cellular contexts, including *ANKRD18A*, *LNCTSI*, *MX1*, *IL10RB*, and *APP* (**Figure 4C**). Forest plots confirmed reproducible overexpression for multiple high-confidence genes, including *DYRK1A*, *IFNAR1*, *IFNAR2*, and *ADARB1* (**Figure 4D–G**). Notably, transcriptome-wide differential expression patterns in our fetal PFC and hippocampus closely mirrored those from independent datasets, underscoring the robustness and cross-context validity of our meta-analytic framework (**Figure S11**).

**Figure 4.**
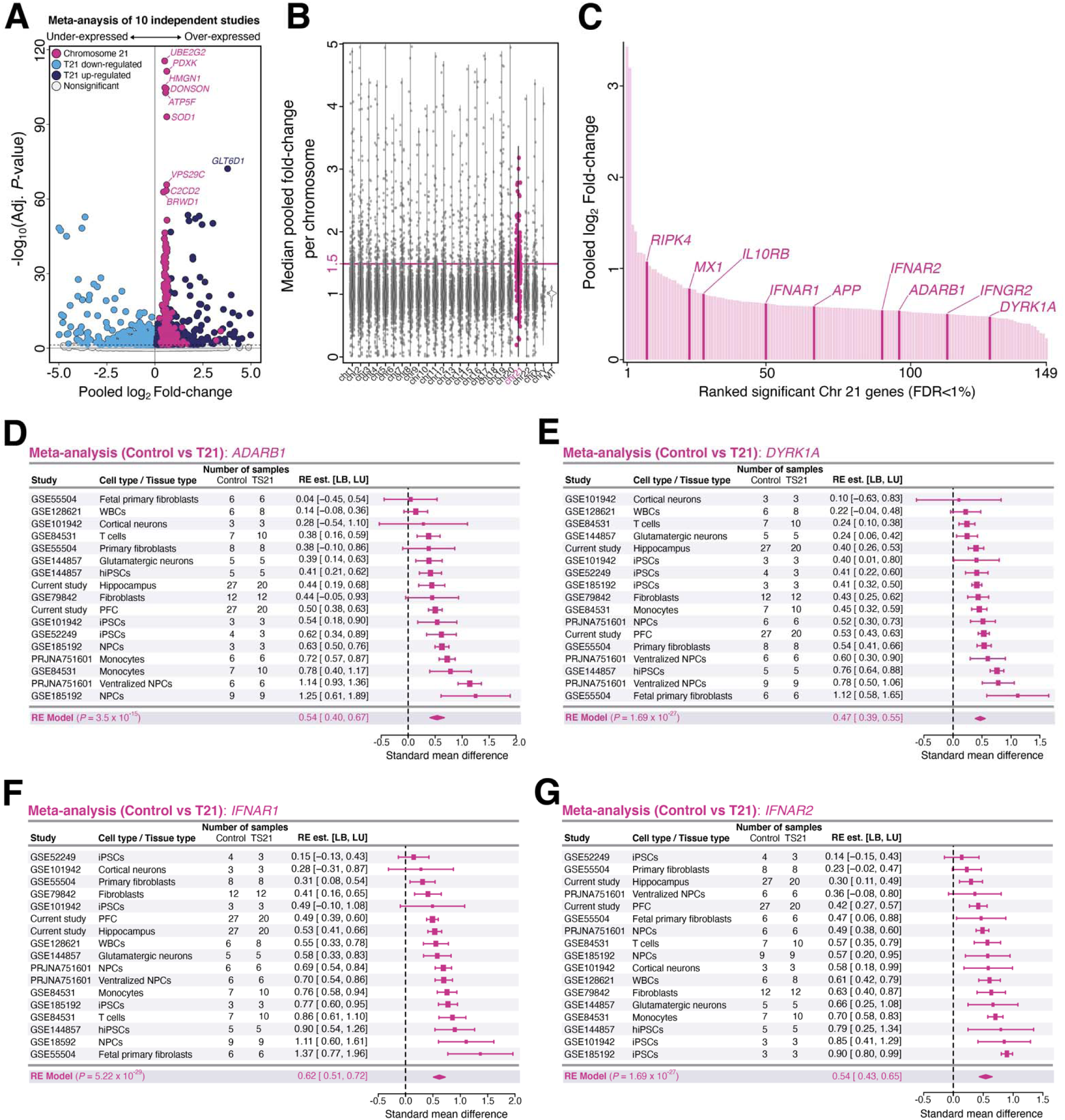
Meta-analysis of gene expression across developmental T21 datasets. (**A**) Volcano plot displaying transcriptome-wide differential expression results from a meta-analysis of ten independent T21 versus control RNA-seq studies. The y-axis indicates statistical significance (−log FDR-adjusted p-value < 1%), and the x-axis reflects log fold-change. Genes are colored according to direction of dysregulation and chromosomal location, with chromosome 21 genes highlighted. The top 10 T21-related over-expressed genes are labeled. (**B**) Average log fold-change (y-axis) per chromosome, showing that chromosome 21 genes approximate the expected ∼1.5-fold increase due to gene triplication. (**C**) Ranked plot of log fold-changes for all significant chromosome 21 genes (FDR <1%), arranged by genomic position (x-axis), illustrating selective rather than uniform dosage sensitivity. Eight well-known T21-related genes and *ADARB1* are highlighted. (**D–G**) Forest plots display standardized mean differences in expression across ten datasets for four high-confidence dosage-sensitive genes: *ADARB1*, *DYRK1A*, *IFNAR1*, and *IFNAR2*. Each plot lists individual datasets on the y-axis, with corresponding effect sizes (dots) and 95% confidence intervals (horizontal lines) plotted along the x-axis. Summary diamonds (pink) denote the combined effect estimates derived from a random-effects meta-analysis, with axes scaled to the minimum and maximum confidence intervals for each gene to facilitate comparison.

Among these, *ADARB1* was consistently overexpressed across most independent studies, making it a particularly compelling candidate for further investigation due to its known role in RNA editing and neurodevelopment. This gene encodes an adenosine deaminase involved in A-to-I editing, a post-transcriptional mechanism that modifies RNA sequences to regulate protein function and neuronal excitability during development9. Despite its consistent overexpression in both our fetal brain dataset and across external studies, *ADARB1* has received relatively limited attention in the context of T21. Its dosage sensitivity, role in RNA editing, and potential interaction with interferon signaling pathways position it as a strong candidate for investigating downstream consequences of gene dosage imbalance. We therefore next examined whether *ADARB1* overexpression in T21 is associated with altered A-to-I editing in the developing brain.

### *ADARB1* is associated with increases in A-to-I editing and excessive RNA recoding

A-to-I editing is catalyzed by the ADAR family of enzymes, including ADAR1 and ADAR2 (*ADARB1*), which act on double-stranded RNA structures to diversify the transcriptome^25^. ADAR1, encoded on chromosome 1, exists in two isoforms: the constitutively expressed nuclear isoform p110 and the cytoplasmic, interferon-inducible isoform p150, which plays a key role in immune signaling^26^. ADAR2 is highly expressed in the central nervous system and mediates site-specific editing of protein-coding transcripts essential for neuronal development^27^. In contrast, ADAR3 (*ADARB2*), also brain-specific, lacks catalytic activity and is thought to act as a dominant-negative regulator of A-to-I editing through competition for RNA substrates^28^.

In our fetal brain RNA-seq data, we observed significant dosage-sensitive upregulation of *ADARB1* in individuals with T21, in both the PFC (FDR *p* = 4.0 × 10 ) and hippocampus (FDR *p* = 0.04), with no significant changes in expression for *ADAR* or *ADARB2* (**Figure 5A, Table S8**). To assess the impact of this dosage effect on global A-to-I editing activity, we computed the *Alu* editing index (AEI), defined as the ratio of A-to-G edited reads to total adenosine coverage in *Alu* elements across the transcriptome (*see Methods*). AEI values were significantly elevated in T21 compared to controls in the PFC (*p* = 0.01, linear regression), and showed a marginal trend in the hippocampus (*p* = 0.05) (**Figure 5B**). To evaluate the contribution of each ADAR enzyme to global editing levels, we performed a bootstrapped linear regression of AEI values against the expression of *ADAR*, *ADARB1*, and *ADARB2*. Among the three, *ADARB1* exhibited the strongest positive association with AEI (bootstrapped slope = 0.27), while *ADAR* (slope = 0.03) and *ADARB2* (slope = 0.11) showed weaker and more variable effects (**Figure 5C**). Confidence intervals for *ADAR* and *ADARB2* were broad, reflecting greater inter-sample variability. These results position *ADARB1* as the primary contributor to elevated A-to-I editing in T21 fetal brain, with the steepest and most consistent slope indicating the greatest per-unit effect of gene expression on editing activity, capturing regulatory influence more directly than correlation-based approaches.

**Figure 5.**
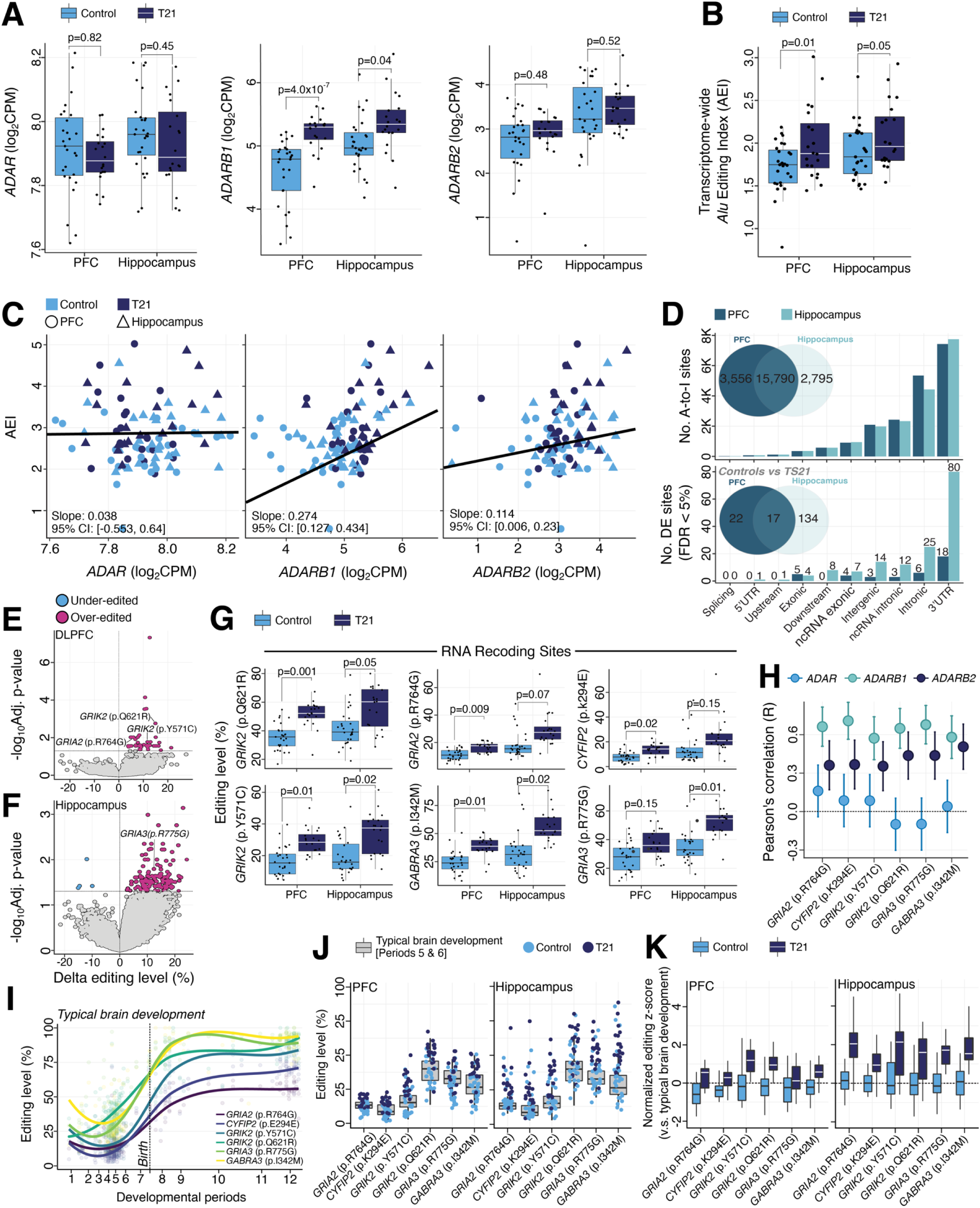
Trisomy 21 is associated with ADARB1 overexpression and increased A-to-I RNA editing activity in the fetal brain. (**A**) Normalized expression levels of *ADAR*, *ADARB1*, and *ADARB2* in the PFC and hippocampus of T21 and control samples. Adjusted *p*-values were computed using moderated t-tests applied transcriptome-wide. (**B**) Alu editing index (AEI), a global measure of A-to-I editing activity, calculated from bulk RNA-seq in PFC and Hippocampus. Significance was assessed by two-sided linear regression. (**C**) Bootstrapped linear regressions showing the relationship between AEI and normalized expression (log CPM) of *ADAR*, *ADARB1*, and *ADARB2*. (**D**) Total number of detected A-to-I editing sites per brain region (top) and the subset identified as differentially edited between T21 and control samples (bottom). Venn diagrams indicate regional overlap in total site detection (top) and differential editing (bottom). Volcano plots depicting changes in editing levels in T21 relative to controls, comparing the strength of significance (-log_10_ Adj. *p*-value; y-axis) to delta editing level (%; axis) for the (**E**) PFC and (**F**) hippocampus. (**G**) Editing level differences between T21 and controls for six functionally characterized RNA recoding sites across the PFC and hippocampus. BH-adjusted p-values reflect moderated t-tests. (**H**) Pearson correlation coefficients between editing levels at the six recoding sites and the expression of each ADAR family member. (**I**) Developmental trajectories of editing levels at the six recoding sites across typical brain maturation, plotted using BrainVar data from prenatal to postnatal stages. (**J**) Overlay of editing levels for T21 and control samples within BrainVar-defined developmental periods 5 and 6. (**K**) Z-score normalization of T21 and control editing levels relative to BrainVar (Periods 5 and 6) means and standard deviations for each site during matched developmental windows, shown for the PFC (left) and hippocampus (right).

We next profiled high-confidence A-to-I sites across the transcriptome using a curated database and a supervised detection pipeline, followed by stringent quality control filtering (*see Methods*). This approach yielded 19,346 sites in the PFC and 18,585 in the hippocampus, with ∼80% of sites shared between regions (**Figure 5D**). As expected, most editing sites were located within 3′UTRs and introns. Next, differential editing analyses were conducted between T21 and control samples, adjusting for estimated neuronal cell type proportions, RIN, gestational age, sex, and percentage of mRNA reads, within each region. We identified 39 dysregulated sites in the PFC and 151 in the hippocampus (FDR < 5%), the vast majority of which were over-edited in T21 (**Figure 5E–F, Table S9**), with predominant localization to 3′UTRs. In fact, 3’UTR sites showed enrichment among differentially edited sites in the PFC (1.20-fold enrichment; *p* = 0.049, one-sided FET) and hippocampus (1.56-fold enrichment; *p* = 1.6 × 10 , one-sided FET). Notably, exonic sites showed significant enrichment for differentially edited sites in the PFC (∼7.1-fold enrichment, *p* = 0.00064, one-sided FET) with a suggestive enrichment in the hippocampus (1.78-fold enrichment; *p* = 0.19, one-sided FET). Across genes, RNA editing in the 3′ UTR showed an inverse correlation with expression (**Figure S12**), consistent with the idea that increased A-to-I editing destabilizes transcripts by altering structure, miRNA binding, or translational efficiency. Similar negative coupling across other genic regions suggests a pervasive link between editing activity and reduced mRNA abundance. Notably, intersecting differentially expressed genes with transcripts harboring over-edited sites revealed minimal overlap, one gene in the PFC (*MRPL30*) and one in the hippocampus (*BASP1-AS1*), indicating limited convergence between transcriptional and editing dysregulation.

Among the differentially edited sites, we identified seven well-characterized RNA recoding events where A-to-I editing results in nonsynonymous amino acid changes that were consistently over-edited in T21 and are known to play critical roles in early brain development (**Figure 5G, Figure S13**). Several of these sites reached statistical significance in both brain regions, while others were significant in one region and showed consistent trends in the other. For example, two recoding sites within *GRIK2* (p.Q621R and p.Y571C) were significantly over-edited in both the PFC (FDR *p* = 0.001, FDR *p* = 0.01, respectively) and hippocampus (FDR *p* = 0.05, FDR *p* = 0.02, respectively). Over-editing was also observed at *GRIA2* (p.R764G) and *GRIA3* (p.R775G) in the PFC (FDR *p* = 0.0009, FDR *p*=0.15, respectively) and hippocampus (FDR *p* = 0.07, FDR *p*=0.01, respectively). Additional recoding sites showing over-editing included *GABRA3* (p.I342M) and *CYFIP2* (p.K294E) in the PFC (FDR *p* = 0.01 and 0.02) and hippocampus (FDR *p* = 0.02 and 0.15), and *COG3* (p.I635V) in the hippocampus (FDR *p* = 0.03) (**Figure S13**). Editing levels at these loci were strongly correlated with *ADARB1* expression (**Figure 5H**), consistent with its known regulatory role in mediating editing at recoding sites.

To further define the molecular identity of T21-associated editing changes, we integrated reference A-to-I editing maps from purified human brain cell types^23^. This analysis revealed that most over-edited sites in T21, particularly those within protein-coding regions, are strongly enriched in neurons, with minimal editing in glial populations (**Figure S14**). This neuron-specific pattern argues against a secondary inflammatory or immune origin. Moreover, cell type deconvolution of bulk RNA-seq showed no significant differences in neuronal composition between T21 and control samples (**Figure S1**), further excluding cell proportion as a confounding factor. Many of these neuron-enriched sites, especially recoding events, were also found to be highly edited in fresh brain tissue but markedly reduced in postmortem samples (**Figure S14**), underscoring their dynamic regulation, and supporting a functional role in neuronal plasticity rather than passive degradation artifacts.

These recoding sites undergo tightly regulated, site-specific changes during typical brain maturation, with editing levels gradually increasing and peaking postnatally, as shown using BrainVar typical cortical developmental data^29^ (**Figure 5I**). To contextualize editing levels in T21, we overlaid our fetal brain data onto these normative trajectories, focusing on BrainVar samples from fetal Periods 5 and 6, which correspond to our gestational window. Across all six selected recoding sites, T21 samples consistently exhibited elevated editing relative to both age-matched controls and the BrainVar developmental distribution (**Figure 5J**). To quantify these shifts relative to typical brain development, we normalized editing levels using z-scores calculated against the BrainVar reference dataset. Specifically, for each site and sample, we computed z-scores using the mean and standard deviation of editing levels from neurotypical BrainVar samples during developmental Periods 5 and 6 (*see Methods*). This normalization anchored our T21 and control measurements to a well-characterized mid-gestational baseline. Across all six sites, T21 samples exhibited consistently positive z-scores, indicating premature or excessive editing relative to normative expectations (**Figure 5K**). In contrast, control samples were centered near the BrainVar developmental mean (**Figure 5K**). These results suggest that T21 is associated with editing at key neurodevelopmental loci, potentially reflecting premature post-transcriptional maturation and altered proteomic remodeling during fetal brain development.

### Meta-analysis of A-to-I editing across diverse tissues confirms over-editing in trisomy 21

Although A-to-I editing is known to vary across cell types, tissues, and developmental stages, we sought to validate our primary findings by performing a cross-study meta-analysis encompassing diverse cellular contexts. Using a harmonized editing quantification pipeline and consistent site detection criteria across datasets, we identified 68 high-confidence A-to-I editing sites under strict FDR <5% and an additional 69 A-to-I sites under FDR <10%, that were over-edited in T21 compared to controls (**Figure 6A**). These sites were selected from a transcriptome-wide screen and detected consistently across multiple studies. ∼51% of these sites (71 sites) localized to 3′ untranslated regions (3′UTRs) (**Figure 6B**), highlighting a potentially conserved role for post-transcriptional dysregulation in T21. Using a harmonized editing quantification pipeline and consistent site detection criteria across datasets, we identified 68 high-confidence A-to-I editing sites that were significantly over-edited in T21 compared to controls at a stringent false discovery rate (FDR < 5%), along with an additional 69 sites reaching FDR < 10% (**Figure 6A**). To complement the high-confidence sites, we expanded the threshold to FDR <10% to cast a broader net and detect consistent, lower-magnitude editing changes that may still be biologically relevant for T21. While more exploratory, these sites were retained only if consistently detected across datasets and offer candidates for future validation (*see Methods*). Notably, ∼51% (71 sites) localized to 3′ untranslated regions (3′UTRs) (**Figure 6B**), underscoring a potential conserved mechanism of post-transcriptional dysregulation in T21.

**Figure 6.**
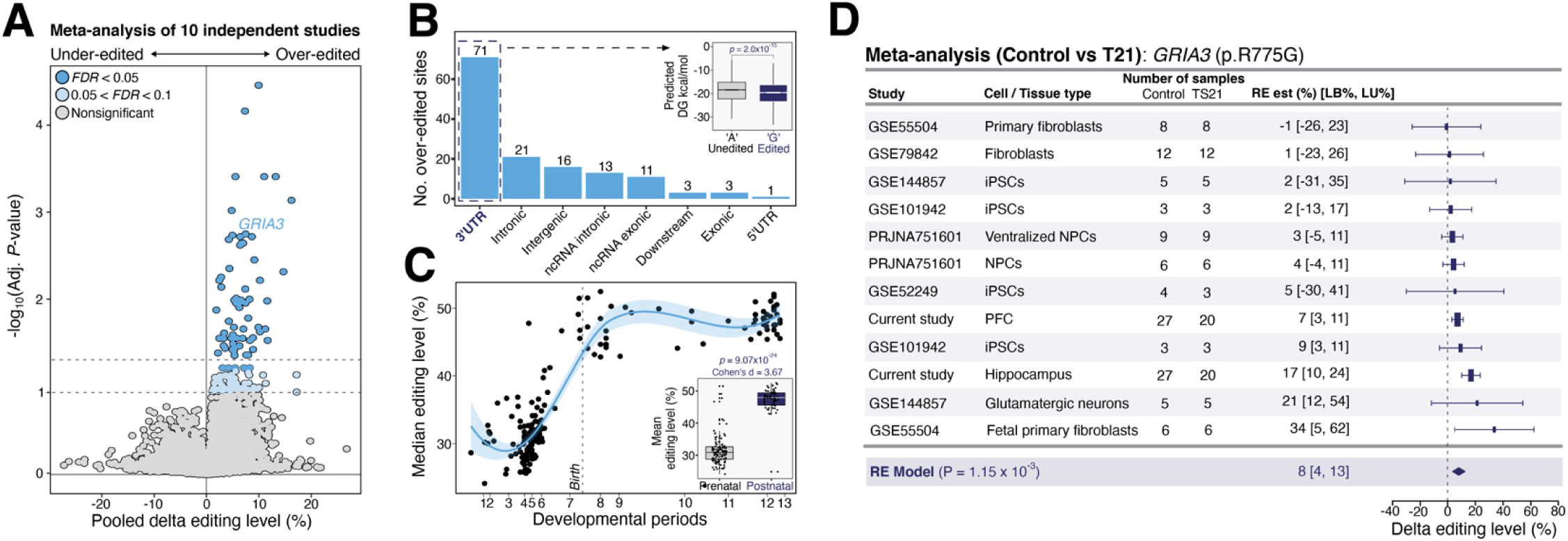
Cross-dataset meta-analysis reveals conserved A-to-I editing dysregulation in trisomy 21. (**A**) Differential RNA editing meta-analysis comparing T21 and control samples across ten independent datasets. Each site is plotted by pooled delta editing level (x-axis) and significance (−log FDR-adjusted p-value; y-axis). Sites are colored by statistical confidence: FDR <1% (red), 5–10% (orange), and >5% (grey). (**B**) Distribution of differentially edited sites by genic region, highlighting strong enrichment in 3′UTRs. Inset: Boxplot comparing miRNA binding affinity (minimum free energy, MFE) between unedited and edited 3′UTRs using high-confidence miRNA seed matches. Lower MFE values reflect stronger predicted binding; significance was assessed using a two-sided Mann–Whitney U test. (**C**) Average editing levels of all over-edited sites in T21 plotted across prenatal to postnatal stages using the BrainVar developmental dataset. Inset: Boxplot showing mean editing levels for these sites in prenatal versus postnatal periods of typical development (p = 9.0 × 10 ² , Cohen’s D = 3.67). (**D**) Forest plot of GRIA3 (p.R775G) editing levels comparing T21 and controls across all datasets. The x-axis shows delta editing as a percentage; black bars denote confidence intervals, and the pink diamond represents the combined random-effects model estimate.

We further modeled the impact of editing on miRNA–mRNA interactions using thermodynamic predictions (*see Methods*). Edited 3′UTRs exhibited significantly lower miRNA binding affinity than their unedited counterparts (*p* = 2.3 × 10 ³², Mann-Whitney U test) (**Figure 6B**), suggesting that over-editing in T21 may disrupt canonical miRNA regulation during critical developmental windows. While these findings are based on computational predictions, they support a mechanistic link between A-to-I editing and post-transcriptional gene silencing in the context of T21.

To further evaluate the developmental relevance of T21 over-edited sites, we examined their editing trajectories across typical cortical development. All sites exhibited increasing editing from the prenatal to postnatal period, with mean levels rising from ∼35% to ∼50% (Cohen’s d= 3.67, **Figure 6C**), suggesting tight temporal regulation during maturation. Among the recoding sites elevated in T21, GRIA3 (p.R775G) emerged as the most consistently over-edited locus across both fetal brain and multiple in vitro models (**Figure 6D**). This site, which regulates AMPA receptor desensitization kinetics, showed a statistically robust ∼8% average increase in editing in T21 (RE model: *p* = 0.0011), with most datasets demonstrating concordant over-editing. Our findings suggest that in T21, premature attainment of these editing thresholds may disrupt synaptic refinement and shift excitatory–inhibitory (E/I) balance during critical periods of circuit formation. Moreover, the reproducibility of this editing phenotype across diverse cell types, from iPSCs and NPCs to fibroblasts and postmortem brain, underscores its biological relevance and supports a model in which *ADARB1*-driven editing at functionally constrained sites contributes to altered excitatory signaling in T21.

### Peripheral blood RNA-seq reveals immune-linked ADAR activation in T21

To contextualize CNS-specific RNA editing dysregulation in T21, we analyzed matched bulk RNA-seq data from peripheral whole blood samples (n=304 T21, n=96 controls)^20^. *ADARB1* expression was significantly elevated in T21 blood (FDR p = 3.14 × 10 ), consistent with gene dosage effects, whereas *ADAR* and *ADARB2* expression were unaltered by karyotype (**Figure S15A**). However, in contrast to the brain, global editing activity (AEI) was not elevated across all T21 samples (**Figure S15B**). To evaluate immune-related editing effects, we stratified samples by transcriptional subtype (MS1–MS3), revealing that only MS3 individuals (characterized by high innate immune activation) exhibited increased AEI and elevated *ADAR* expression (**Figure S15C–D**). Notably, *ADARB1* expression tracked with karyotype but not immune tone and did not correlate with AEI (*r* = –0.0514, *p* = 0.24), whereas *ADAR* strongly correlated with AEI (*r* = 0.599, *p* = 1.07 × 10 ³) (**Figure S15E**). These findings highlight that while *ADARB1* is dosage-sensitive in both tissues, its role in RNA editing is CNS-specific, whereas *ADAR* mediates immune-linked editing in peripheral blood. This contrast underscores tissue-specific regulatory mechanisms and supports a dual model in which gene dosage and immune signaling independently shape A-to-I editing dynamics across biological systems.

## DISCUSSION

Despite the well-established genetic basis of DS as trisomy of chromosome 21, the mechanisms by which this chromosomal imbalance disrupts early human brain development remain poorly understood. Prior transcriptomic studies in fetal and postnatal tissues have reported widespread gene expression changes, implicating both direct dosage effects of chromosome 21 genes and secondary perturbations in neurodevelopmental, glial, and immune signaling pathways^2–7^. However, these studies have often been constrained by limited sample sizes, incomplete transcriptome survey, postnatal sampling windows, or insufficient cell type resolution. To address these gaps, we integrated bulk RNA-seq data from the PFC and hippocampus of mid-gestation T21 fetal brains with a harmonized meta-analysis of nine additional independent RNA-seq datasets spanning early diverse T21 cellular contexts (**Figure 1**, **Table 1**). Our findings highlight A-to-I editing, particularly through *ADARB1* overexpression, as a conserved, dosage-sensitive mechanism of transcriptomic dysregulation in T21. By linking increased editing to both coding and regulatory RNA elements, this study identifies RNA editing as a previously underappreciated axis of gene dysregulation during fetal brain development. Notably, these effects converge across cell types, brain regions, and independent datasets, highlighting RNA editing as a cross-cutting mechanism that integrates gene dosage imbalance with altered post-transcriptional regulation. Given the timing and sensitivity of *ADARB1*-driven editing, these findings raise the possibility that RNA editing profiles may serve as early biomarkers of circuit maturation and suggest new therapeutic entry points for restoring post-transcriptional balance in DS. Importantly, this work reframes T21 neuropathology through the lens of dynamic RNA regulation, complementing existing transcriptomic and epigenomic models with a mechanism tightly linked to developmental timing and neuronal function. To facilitate further research, all data from this study are available via an open-access, interactive online resource (*see Data Availability*).

Our transcriptomic analysis revealed gene dysregulation across fetal PFC and hippocampus in T21, including a pronounced enrichment of chromosome 21 genes (**Figure 2**). While previous studies reported variability in chromosome 21 dosage effects, ranging from global overexpression to partial compensation^7,18,20,30,31^, our data reinforce a model of selective dosage sensitivity. Specifically, chromosome 21 genes that are lowly expressed in the fetal brain are more likely to escape significant upregulation in T21, suggesting that transcriptional buffering or tighter regulatory control may limit their dosage response. This implies that gene expression changes in T21 are not uniformly driven by copy number, but instead shaped by context-dependent regulatory mechanisms, reinforcing a non-global model of gene dosage effects. Genes such as *DYRK1A*, *IFNAR1*, and *IFNAR2* were consistently overexpressed, whereas others appeared transcriptionally buffered. Beyond chromosome 21, downregulated genes were enriched for mitochondrial translation and ribosomal biogenesis, indicative of impaired metabolic capacity during neurogenesis^32,33^. In contrast, upregulated non-chromosome 21 transcripts, including *DCN* and *LTBP4*, suggested atypical extracellular matrix remodeling and early electrophysiological maturation^3,34^. Permutation-based gene set preservation analysis revealed marked disruption of gene regulatory architecture in the hippocampus, particularly in modules governing synaptic plasticity, chromatin remodeling, and glial immune signaling (**Figure 3**), suggesting region-specific vulnerability to dosage imbalance and its downstream effects.

To assess the generalizability of these patterns and to synthesize extant data, we performed a harmonized meta-analysis, integrating our fetal RNA-seq data with 9 independent RNA-seq datasets from T21 cases and controls (**Figure 4**). This analysis yielded a robust signature of 543 upregulated and 437 downregulated genes (FDR < 1%), of which 30.7% of the upregulated genes were chromosome 21 encoded, closely mirroring the expected 1.5× gene dosage effects. However, consistent with our fetal brain findings and previous analyses, only a subset of chromosome 21 genes exhibited reproducible overexpression^,18,20,30,31^. Among these, *ADARB1* emerged as one of the most consistently upregulated genes across brain and non-brain tissues. In T21 fetal brains, *ADARB1* was significantly elevated in both the PFC and hippocampus, while expression of other ADAR family members (*ADAR1*, *ADARB2*) remained unchanged. This association is robust and consistent across datasets and supported by independent A-to-I editing profiles in the human brain^16,28,29,35^. Our findings also support observations from previous *in vitro* studies using T21 iPSCs^13^, which noted *ADARB1* dosage increases and potential impacts on editing fidelity and transcript diversity. While Kawahara et al. reported no changes in *ADAR2* expression or GRIA2 Q/R site editing in one neonate DS brain tissue^12^, our brain-anchored data suggest that sample size, developmental timing and/or regional specificity may reveal editing dysregulation not captured in whole-tissue analyses.

Transcriptome-wide analyses revealed consistent over-editing at a subset of A-to-I sites, including seven protein-coding recoding events critical for neuronal signaling^8,9^ **(Figure 5, Figure S13**). These included the Q/R site in *GRIA2*, as well as homologous sites in *GRIA3* and *GRIK2*, all of which undergo tightly regulated, developmentally timed editing that governs calcium permeability and receptor desensitization kinetics^36,37^. Among the consistently over-edited sites in T21 fetal brain, the GRIA3 R/G site (p.R775G) showed a ∼8% increase in editing across brain regions and in vitro models (**Figure 6**). Although editing at this site is conserved and has been implicated in regulating AMPA receptor desensitization kinetics^38,39^, its *in vivo* functional significance remains incompletely characterized. In contrast to the well-studied GRIA2 Q/R site, whose loss of editing accounts for the severe phenotypes in Adar2 knockout mice^40–42^, there is currently no direct evidence linking GRIA3 R/G editing to developmental phenotypes *in vivo*. However, increased GRIA3 editing has been observed in the context of epilepsy^43,44^, suggesting that it may respond to or modulate excitatory signaling. Furthermore, ADAR2 overexpression in rodents has been associated with behavioral phenotypes such as hyperphagia^45^, although these phenotypes have not been causally linked to GRIA3 editing or to T21. While we observe a robust and premature shift in editing at GRIA3 R/G and other recoding sites, we interpret these findings as correlative with *ADARB1* upregulation rather than directly causal. Future site-directed editing studies will be needed to determine whether specific recoding changes contribute to altered neurodevelopment in T21.

Additional over-edited transcripts included *GABRA3* (I342M), which modulates GABA_A receptor gating and trafficking, and *CYFIP2* (K294E), a synaptic scaffolding protein involved in dendritic spine remodeling and local translation^46,47^. These alterations likely compound the effects of glutamate receptor dysregulation and disrupt synaptogenesis. Supporting this view, DS-derived astrocytes have been shown to impair synapse formation and induce mTOR hyperactivation in co-cultured neurons, a phenotype linked to E/I imbalance and reduced connectivity^48^. This is consistent with recent findings demonstrating that T21 iPSC-derived astrocytes secrete factors that impair neuronal mTOR signaling and reduce synaptic density *in vitro*^49^. These non-cell-autonomous effects may act synergistically with post-transcriptional disruptions, such as premature RNA editing, to alter receptor composition and shift the developmental timing of synaptic integration. Moreover, these recoding sites are also among the most highly edited in living (antemortem) human brain tissue but are markedly less edited in postmortem samples (**Figure S14**)^23^, highlighting their reliance on intact cellular physiology. Their premature over-editing in T21 may therefore reflect a shift in the temporal regulation of editing normally restricted to viable, active neural circuits, further supporting their functional relevance during early synaptogenesis

To contextualize the CNS-specificity of our findings, we extended our analysis to peripheral blood RNA-seq data from individuals with T21 and controls^20^. *ADARB1* expression was significantly elevated in T21 blood samples, consistent with gene dosage effects, yet global editing activity (AEI) was not increased (**Figure S15**). However, when stratified by transcriptional subtype, only individuals with high interferon signaling (MS3 group) exhibited elevated AEI alongside increased *ADAR* expression. These observations underscore canonical immune-linked editing dynamics in blood, in contrast to the fetal brain, where AEI was driven by *ADARB1* expression, independent of immune activation or *ADAR* levels. This divergence highlights a tissue-specific regulatory axis in T21: *ADARB1* predominantly governs CNS editing, consistent with our prior reports^16,28,29,35^, while *ADAR* responds to inflammatory signaling in immune tissue^21,22^. Notably, we also observed a positive correlation between *ADARB2* and AEI in brain tissue. Despite its lack of catalytic activity, *ADARB2* may act as a dominant-negative modulator or reflect co-regulated expression patterns with ADARB1 during development. Together, these findings support a dual regulatory model of RNA editing in T21, shaped by both gene dosage and immune signaling, and emphasize the importance of tissue context in interpreting A-to-I editing dynamics.

Several limitations of this study should be acknowledged. First, while bulk RNA-seq offers robust insight into global transcriptomic changes, it lacks the resolution to resolve cell type–specific editing patterns. Future studies leveraging single-nucleus or spatial transcriptomics will be essential to map editing dynamics across distinct neuronal and glial populations. Second, although our meta-analysis employed a harmonized processing pipeline, heterogeneity in source datasets, including differences in cell type composition, developmental stage, and technical protocols, may have reduced sensitivity for detecting low-abundance or cell type-specific editing events. To mitigate this, we prioritized the identification of T21-associated editing changes that were consistently observed across diverse cellular contexts. Finally, while the observed association between *ADARB1* and aberrant editing is compelling, causal relationships remain to be established. For example, we did not perform functional assays to test the impact of individual recoding sites such as *GRIA3* R/G. Experimental perturbation of individual recoding events, such as those affecting *GRIA3* or 3′UTR-miRNA interactions, are required to determine their precise contribution to T21-associated phenotypes.

In sum, this study identifies RNA editing as a novel, dosage-sensitive layer of transcriptomic dysregulation in T21, driven in part by the overexpression of *ADARB1*. This finding extends prior work on the dynamic regulation of A-to-I editing across brain development, where precise timing and spatial control are essential for synaptic homeostasis and neuronal plasticity. Here, we show that *ADARB1-*driven editing dysregulation emerges earlier than previously recognized, impacting a distinct set of neurodevelopmental targets. By integrating fetal brain sequencing, gene co-expression analysis, and cross-system meta-analysis, we define a reproducible molecular signature linking chromosome 21 dosage to post-transcriptional disruption. These findings position RNA editing as a regulatory amplifier of T21’s molecular effects, with the potential to exacerbate synaptic dysfunction. As such, RNA editing signatures may serve as biomarkers of circuit maturation and offer novel entry points for correcting post-transcriptional imbalance in DS.

## METHODS

### Postmortem Tissue Acquisition and DNA Testing

All tissue samples were collected at University Hospital of Obstetrics and Gynecology, Sofia, in collaboration between Lieber Institute for Brain Development (LIBD) and Medical University of Sofia, Bulgaria. The postmortem brain tissues were collected after written informed consent was obtained from the mothers and stored fresh frozen at the biobank of Molecular Medicine Center, Medical University of Sofia before transport to LIBD for further processing. Altogether fetal brain tissues from 20 T21 cases and 27 euploid controls between 13–22 gestational week were used in the study. All 20 T21 cases analyzed in this study were confirmed as full trisomy 21 by clinical karyotyping reports obtained from the source biobank. No partial or mosaic cases were included. The fetal material was collected after termination of pregnancy for medical reasons (severe congenital anomalies or genetic disorder). Invasive prenatal aneuploidy testing using Aneufast™ QF-PCR Kit (Molgentix Sl) was performed at the University Hospital of Obstetrics and Gynecology and microarray testing of DNA from the abortive tissue was done using SurePrint G3 Human CGH Microarray Kit, 4 × 180K (Agilent Technologies Inc) at Molecular Medicine Center. The research project “Genetic and epigenetic research of human fetal brain development” was approved by the Institutional Ethics Review Board of Medical University of Sofia, Protocol No. 2981/06.07.2012.

### RNA-sequencing data generation

We assembled a discovery cohort consisting of paired prefrontal cortex (PFC) and hippocampal formation tissue samples (referred to as “hippocampus” throughout this body of work) from 20 individuals with trisomy 21 (T21) and 27 euploid controls, collected between 13 and 22 post-conception weeks (PCW), a critical period for human brain development. At these early developmental stages, the hippocampal formation represents the broader precursor region giving rise to the mature hippocampus. Donor metadata are detailed in **Table S1**. The cohort included 16 females and 31 males, with a mean gestational age of 18.4 ± 3.2 weeks. All samples were from individuals of Caucasian ancestry with no reported major brain malformations. Total RNA was extracted from frozen tissue using the AllPrep DNA/RNA Mini Kit (Qiagen, Cat No. 80204). RNA quality was assessed using the Agilent 2100 Bioanalyzer; all included samples had RNA Integrity Numbers (RIN) ≥ 9.0 (9.8 ± 0.3). RNA-seq libraries were prepared from 300 ng of total RNA using the TruSeq Stranded Total RNA Library Prep Kit with Ribo-Zero Gold (Illumina), which depletes both cytoplasmic and mitochondrial ribosomal RNA. Libraries were sequenced on an Illumina HiSeq 3000 platform at the Promega sequencing core, generating 200-bp paired-end reads at an average depth of ∼34.5 million reads per sample.

### RNA-sequencing data processing and quality control

Sequencing reads were processed using the RAPiD-nf pipeline (https://github.com/CommonMindConsortium/RAPiD-nf). Reads were aligned to the GRCh38 human reference genome (Ensembl release 95) using STAR v2.7.3a^50^. Alignment quality was assessed using RSeQC and Picard tools, including metrics for GC content, duplication rates, insert size, gene body coverage, and library complexity. PCR duplicates were marked but retained during quantification. Gene-level quantification was performed using featureCounts^51^ (Subread v2.0.1) based on GENCODE v32 annotations. Genes with non-zero counts in at least one-third of all samples were retained. Counts were normalized using the VOOM transformation from the limma package (v3.48.3)^52^ to form region-specific expression matrices (PFC and hippocampus). A total of 16,472 genes were analyzed in the PFC and 17,035 genes were analyzed in the hippocampus. Principal component analysis (PCA) did not identify any expression outliers using a 95% confidence interval across the top three PCs.

### Cellular deconvolution of bulk tissues

Cell-type composition was estimated using non-negative least squares (NNLS) implemented in the est_frac function of the bMIND R package (v1.2.0) ^53^. This approach applies Bayesian inference to estimate cell-type–specific expression profiles while constraining coefficients to non-negative fractions. Prior information for the model was derived from a reference single-cell expression signature matrix from Pei et al. (2021) ^54^. This matrix includes transcriptional profiles for 11 fetal brain cell types: astrocytes, microglia, oligodendrocytes, oligodendrocyte precursor cells (OPCs), endothelial cells, pericytes, intermediate progenitor cells (IPCs), neuroepithelial cells (NEPs), and three neuronal subtypes (quiescent, replicating, transitional). Log2-transformed counts per million (CPM) were used as input^55^. Deconvolution results were used to adjust for cellular heterogeneity in downstream modeling. This approach enabled us to assess and adjust for potential shifts in cell type composition while minimizing confounding due to cellular heterogeneity, consistent with prior studies of fetal brain transcriptomics.

### Partitioning gene expression variance

We used the variancePartition R package^56^ to fit linear mixed models quantifying the proportion of expression variance attributable to technical and biological factors. Covariates included neuronal proportion (quiescent + replicating + transitional neurons), % mRNA reads, RIN, gestational age, sex, library depth, and genetic diagnosis. Donor identity was modeled as a random effect. These estimates informed covariate selection for differential expression and network modeling.

### Differential gene expression analysis

Differential expression was performed using the limma-voom pipeline^55^. An initial analysis compared PFC versus hippocampus, adjusting for diagnosis, percentage of mRNA reads, gestational age, sex, and RIN, with donor modeled via duplicateCorrelation. T21-specific effects were then assessed separately for each region using region-specific VOOM-normalized matrices (16,471 genes in PFC; 17,035 in hippocampus). Models included covariates for neuronal proportion, percentage of mRNA reads, age, sex, and RIN. DEGs were defined as those with Benjamini-Hochberg FDR < 0.05.

### Weighted Gene Co-expression Network Analysis (WGCNA)

To identify coordinated transcriptional programs disrupted in T21, we performed weighted gene co-expression network analysis (WGCNA)^57^ separately for the PFC and hippocampus using VOOM-normalized expression matrices. A total of 16,471 genes (PFC) and 17,035 genes (hippocampus) were retained. Signed co-expression networks were constructed by computing absolute Pearson correlation coefficients between all gene pairs, followed by soft-thresholding to approximate scale-free topology (target R² > 0.80). Power parameters (β) of 5 and 9 were selected for the PFC and hippocampus, respectively, based on scale-free fit and mean connectivity criteria. Networks were clustered using the dynamic tree cut algorithm with a minimum module size of 50 and cut height of 0.999. Modules were summarized by their eigengenes (first principal component of each module’s expression matrix) and tested for association with T21 diagnosis using linear models in limma, adjusting for the same covariates as in the differential gene expression analyses (neuronal proportion, % mRNA, age, sex, and RIN). Module significance was assessed via moderated *t*-tests. Qualitative inspection of module eigengene expression across diagnostic groups was used to confirm biologically interpretable patterns. These analyses enabled the identification of gene networks associated with T21 that may not be captured through single-gene differential expression alone.

### Gene Ontology and Enrichment Analysis

To interpret the functional significance of differentially expressed genes, we performed gene set enrichment analysis using the CAMERA method^58^, a correlation-adjusted competitive test that accounts for inter-gene correlation within gene sets. Differential expression t-statistics were input from the limma-based comparisons between T21 and control samples for each brain region. CAMERA ranks genes based on differential expression and tests whether predefined gene sets are overrepresented at the extremes of this ranked list relative to random expectation. Gene sets were derived from Gene Ontology (GO) biological processes, molecular functions, and MSigDB hallmark gene sets. Variance inflation due to inter-gene correlation was corrected using the default inter-gene correlation parameter (set to 0.01) and resulting enrichment *p*-values were adjusted for multiple testing using the Benjamini-Hochberg method. Enrichment results were used to identify biological processes and pathways disproportionately affected in T21 fetal brain tissues.

### Permutation-Based Gene Set Preservation in T21

We assessed whether co-regulatory structures within predefined gene sets were disrupted in T21 using the Zsummary preservation statistic from WGCNA^59^, computed with 200 permutations. This statistic integrates measures of intramodular density and connectivity across conditions. Zsummary scores <2 indicate no preservation, 2–10 suggest weak to moderate preservation, and >10 reflect strong preservation. Because unsupervised gene co-expression network structure appeared preserved between T21 and controls, we focused on smaller curated gene sets to detect more targeted disruptions, including GO biological processes, GO molecular functions, and MSigDB hallmark gene sets. To correct for the known dependency of preservation scores on module size, we modeled Zsummary of a random geneset matched for geneset size, as a comparative for each tested geneset, as a function of module size and derived an expected preservation score (Zrandom_predicted) for each gene set. We then computed an adjusted rank score incorporating (1) the observed Zsummary, (2) its FDR-adjusted p-value, and (3) the deviation of random module preservation from expectation. Gene sets meeting strict criteria (Zsummary< 2, p.adj > 0.1, Zsummary ≤ Zrandom_predicted− 3, and p.random.adj < 0.05) were classified as non-preserved. These modules exhibited significantly lower preservation than size-matched random sets (Wilcoxon p < 0.05, PFC and hippocampus), supporting loss of co-regulatory structure in biological pathways.

### Cross-Study Meta-Analysis of RNA-Seq Datasets

To validate and generalize transcriptomic findings across diverse developmental contexts, we performed a harmonized meta-analysis of our data with nine additional publicly available RNA-seq datasets profiling T21 and euploid control samples (**Table 1**). These datasets spanned a range of early gestational tissues and in vitro cell types, including induced pluripotent stem cells (iPSCs), neural progenitor cells (NPCs), cortical neurons, and fetal fibroblasts and immune cells. In total, the meta-analysis encompassed 108 T21 and 117 control samples across the following GEO accession numbers: GSE52249, GSE185192, PRJNA751601, GSE101942, GSE144857, GSE55504, GSE79842, GSE84531 and GSE128621. For each dataset, raw FASTQ files were reprocessed through a standardized RNA-seq pipeline (RAPiD-nf; https://github.com/CommonMindConsortium/RAPiD-nf) to ensure uniformity in alignment, quantification, and normalization. Reads were aligned to the human genome (hg38), and gene-level expression was quantified using featureCounts^51^. Differential gene expression within each dataset was computed using limma with voom transformation^46^, adjusting for relevant covariates such as age, sex, batch, and neuronal fraction where available.

For each gene, log fold-change and standard error estimates were extracted from limma-voom analyses of each dataset. Genes with low expression (not expressed in at least one-third of samples per dataset) were excluded. Meta-analysis was performed using a random-effects model implemented via the rma.uni() function in the metafor R package (REML estimator) was used to account for between-study heterogeneity^60^. Only genes expressed in ≥4 datasets and with consistent directionality in at least 70% of datasets were retained in the final result set. Genes with Benjamini-Hochberg adjusted p-values < 0.05 were considered significantly dysregulated. Meta-analysis summary statistics, including forest plots for key dosage-sensitive genes, are publicly available (https://andyyang.shinyapps.io/Ts21-dashboard/). This approach enabled robust quantification of consistent transcriptional changes across diverse developmental contexts and experimental platforms.

### T21 Microarray Data Reprocessing and Differential Expression Analysis

To evaluate the comparability of our RNA-seq findings with prior transcriptomic data, we reprocessed and analyzed a publicly available fetal brain microarray dataset profiling Trisomy 21 (T21) and control samples (GEO accession: GSE59630)^3^ This dataset was generated using the Affymetrix Human Exon 1.0 ST Array platform and includes 58 paired tissue samples from 15 T21 and 15 euploid control brains ranging in age from 14 post-conception weeks (PCW) to 42 years old. For consistency with our fetal RNA-seq dataset, we focused our analysis on the cerebral cortex and hippocampus, corresponding to the neocortical (CBC) and hippocampal (HIP) samples in the original study.

Raw .CEL files were downloaded and imported using the oligo package in R^61^. Robust Multi-array Average (RMA) normalization^62^ was applied to compute expression values at the exon level, followed by log transformation. Metadata for all samples was manually curated and aligned to expression profiles, with key covariates including sex, post-mortem interval (PMI), RNA integrity number (RIN), ethnicity, anatomical region, and developmental stage (converted to numeric age in years, accounting for fetal and postnatal time points). Genes were filtered to retain those expressed above the sample-wise median in at least one-third of samples, resulting in a matrix of robustly expressed transcripts. Gene expression variance was decomposed using the variancePartition R package^56^, employing a linear mixed model that included fixed effects for age, RIN, PMI, and developmental period, and random intercepts for sex, ethnicity, and diagnosis. Differential expression was assessed using the limma framework. A design matrix was constructed to model expression as a function of T21 diagnosis, RIN, PMI, age (in years), and developmental period. Duplicate samples per subject were accounted for by controlling for correlated measurements. A contrast was specified to test for differential expression between DS and control samples. Empirical Bayes moderation was applied to stabilize variance estimates. Significance was defined using a Benjamini-Hochberg false discovery rate (FDR) threshold of <1%.

Fold-changes were directionally aligned to match RNA-seq comparisons, with log fold-changes sign-inverted such that positive values indicate higher expression in T21. Differentially expressed genes (DEGs) were classified as upregulated in T21, upregulated in controls, or non-significant. The resulting gene-level summary statistics were exported for comparison with RNA-seq–derived DEG lists.

### RNA Editing Site Detection and Annotation

To quantify RNA editing across fetal brain samples, we employed a supervised approach that focused on known A-to-I editing sites. Specifically, we queried nucleotide positions curated from high-confidence databases: (i) REDIportal^63^, (ii) previously annotated sites from human brain cell types^28^, and (iii) a comprehensive list of evolutionarily conserved A-to-I recoding sites^64^. Using samtools mpileup, we extracted reference and edited read counts for these known sites directly from STAR-aligned BAM files, following our previously validated pipelines^16,28,29,35^. We then applied stringent quality control criteria to ensure the inclusion of bona fide A-to-I editing events. Supervised sites with <5 total reads or <3 edited reads were excluded. The resulting set of high confidence editing sites was functionally annotated using ANNOVAR^65^, assigning gene symbols based on RefGene, repetitive elements based on RepeatMasker v4.1.1^66^, and known editing annotations via the latest REDIportal database. Evolutionary conservation scores were incorporated using phastCons scores from the PHAST package^67^ to highlight conserved editing events across mammals.

### Alu Editing Index (AEI)

To measure global RNA editing activity, we calculated the *Alu* Editing Index (AEI)^68^, a quantitative metric reflecting A-to-I editing levels in repetitive SINE/Alu elements, where the majority of non-coding editing occurs. The AEI was computed using AEI v1.0, which takes STAR-aligned BAM files as input. Specifically, the AEI is calculated as the percentage of A-to-G mismatches relative to total adenosine coverage within annotated Alu elements (from UCSC RepeatMasker annotations, hg38). Known polymorphic sites were masked using coordinates from the UCSC CommonGenomicSNPs150 track to reduce false positives due to DNA variation. The AEI method preserves signal even in low-coverage regions and has demonstrated robustness across hundreds of postmortem human brain RNA-seq samples generated with varied protocols. AEI values were multiplied by 100 to report editing levels as percentages. This metric served as a transcriptome-wide surrogate for ADAR-mediated editing activity and was used to assess global dysregulation in T21 across brain regions and external replication datasets.

### Quantifying and Normalizing RNA Recoding Dynamics in Typical and Trisomy 21 Fetal Brain

To assess RNA recoding dynamics during fetal brain development and in trisomy 21 (T21), we analyzed A-to-I editing levels at canonical recoding sites across two datasets: (1) neurotypical mid-fetal cortical samples (Periods 5 and 6) from the BrainVar project chromosome 21, and (2) fetal PFC samples from individuals with T21 and matched controls (this study). Editing levels were computed as the ratio of guanosine (G) reads to total adenosine + guanosine (A+G) reads at each site, based on samtools pileups. To contextualize editing disruptions in T21, we visualized recoding site editing levels in T21 and control samples against the backdrop of typical developmental trajectories. Specifically, for each recoding site, we generated boxplots using BrainSpan Period 5–6 PFC samples and overlaid individual T21 and euploid fetal brain samples as points. This approach allowed for direct visual comparison of editing levels relative to normative developmental distributions.

To formally quantify deviation from typical editing patterns, we computed z-scores for each recoding site and individual sample. Z-scores were calculated using the mean (μ) and standard deviation (σ) of editing levels in typical brain development (TBD) BrainVar samples (Periods 5–6, PFC only), according to:

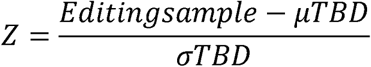

This yielded a normalized deviation score for each site per sample, allowing quantitative comparison across groups. Z-scores were visualized separately by diagnosis (T21 vs. control) and brain region. Group differences in z-scores were assessed using two-sided Wilcoxon rank-sum tests. To account for multiple comparisons across recoding sites, p-values were adjusted using the Benjamini-Hochberg procedure (FDR < 0.05 threshold). This approach enabled the identification of recoding events where editing in T21 significantly deviated from neurotypical developmental trajectories.

### RNA Editing Meta-Analysis Across Developmental Datasets

For each editing site, the mean difference in editing rate (T21 – control) and corresponding standard error were computed from each dataset using linear models adjusted for relevant covariates when provided in the corresponding metadata files (e.g., age, batch, RNA quality). Sites with low coverage (<10 total reads in at least one-third of samples) were excluded to ensure reliability. Editing site meta-analysis was performed using a random-effects model implemented via the rma.uni() function in the *metafor* R package, using the restricted maximum likelihood (REML) estimator to account for between-study heterogeneity. Only sites detected in ≥4 datasets and exhibiting consistent directionality of effect (same sign in at least 70% of datasets) were retained. Sites with Benjamini-Hochberg adjusted p-values < 0.05 were considered significantly differentially edited. This analysis identified high-confidence A-to-I sites consistently over-edited in T21 across fetal brain and in vitro models.

### miRNA-mRNA interactions

miRNA-mRNA interactions were studied using miRANDA^69^ as previously described^29, 70^. In brief, to assess the impact of RNA editing on miRNA binding, we compared the minimum free energy (ΔG) of miRNA-3′UTR interactions at edited versus unedited sites. Next, we extracted matched 101 bp sequences surrounding each editing site (with either A or G), aligned them to all mature miRNAs using miRANDA, and calculated ΔG for each interaction. focusing especially on seed region matches using strict parameters. Statistical differences in ΔG were evaluated using the Mann-Whitney U test.

### Secondary T21 Peripheral blood RNA-sequencing data analysis

To examine ADAR expression and A-to-I RNA editing profiles in peripheral blood, we analyzed bulk RNA-seq data from 304 individuals with T21 and 96 euploid controls obtained from GEO (GSE190125)^20^. Raw FASTQ files were processed using the same standardized pipeline as for the fetal brain and in vitro datasets to ensure analytic consistency. Reads were aligned to the GRCh38 reference genome using STAR^50^, and gene-level expression quantification was performed with featureCounts^51^. *ADAR*, *ADARB1*, and *ADARB2* expression levels were extracted from VOOM normalized^46^ values for downstream analysis. The AEI^68^ was calculated for each sample as a transcriptome-wide proxy for global editing activity. To investigate subgroup-specific effects, we stratified individuals by previously defined transcriptional subtypes from the original study (e.g., MS1, MS2, MS3). Differences in gene expression between T21 and controls, and among molecular subgroups, were performed using limma. All analyses were conducted in R (v4.2.2).

### Data availability

To enable gene-by-gene inspection of between-study effect sizes and heterogeneity, we developed an interactive RShiny App ([https://andyyang.shinyapps.io/Ts21-dashboard/]) that generates meta-analytic forest plots for any gene of interest across the ten harmonized datasets. For each gene, this tool displays study-specific log fold changes (LogFC), standard error, standardized mean differences (SMD), confidence intervals, and random-effects model estimates. This platform provides transparent access to between-study variability, strengthens confidence in meta-analysis results, and enables users to identify genes with consistent or context-specific dysregulation across tissues, cell types, and developmental stages. In addition to meta-analytic forest plots, the app provides dynamic visualizations for each gene: expression boxplots comparing cases and controls, temporal expression trajectories during early human brain development, and normative expression profiles across brain regions using data from BrainSpan. These features enhance interpretability of transcriptomic signals by revealing gene-specific patterns of dysregulation and developmental timing, while enabling transparent access to between-study variability and tissue-specific effects. All original RNA-sequencing data are publicly at the National Center for Biotechnology Information Gene Expression Omnibus under the following accession number: GSE301886.

### Code availability

Computational code is provided on GitHub (https://github.com/BreenMS) and includes R code for: RNA-sequencing quality control, short-read mapping, counting, and normalization, differential gene expression, cell type estimation, A-to-I editing quantification, differential A-to-I editing comparisons, and meta-analyses. Contact the lead author (M.S.B) for any questions.

## Data Availability

https://www.ncbi.nlm.nih.gov/geo/query/acc.cgi?acc=GSE301886

## Acknowledgements

We sincerely thank the families of the brain donors for their invaluable contributions to this research. Without their generosity and willingness to support scientific discovery, this work would not have been possible. We also acknowledge the Lieber Institute for Brain Development Human Brain Repository and collaborating institutions for providing access to these precious samples.

## Funding

This work was supported by the Beatrice and Samuel A. Seaver Foundation. MSB is a Seaver Foundation Faculty Scholar. This work was also supported by the Lieber Institute for Brain Development with funding provided by the families of Constance and Steven Lieber and Milton and Tamar Maltz.

## Competing Interest Statement

DRW serves on the advisory board of Pasithea Therapeutics. All other authors have no competing interests.

## Notes

### Author Declarations

The research project Genetic and epigenetic research of human fetal brain development was approved by the Institutional Ethics Review Board of Medical University of Sofia, Protocol No. 2981/06.07.2012.

