## Supplemental Figures 1-15 for "Trisomy 21 Drives *ADARB1* Overexpression and Premature RNA Recoding in the Developing Fetal Brain"

[*] Shared first authorship
[†] Co-corresponding authors

**Summary of Supplemental Figures**

**Figure S1.** Cell type deconvolution of fetal brain RNA-seq data.
**Figure S2**. Quality control of normalized fetal brain RNA-seq data.
**Figure S3**. Differential gene expression between anatomical regions.
**Figure S4.** Variance of gene expression explained within each region.
**Figure S5**. Unsupervised clustering of differential gene expression signatures.
**Figure S6**. Expression levels of chromosome 21 sensitive and non-sensitive genes.
**Figure S7**. WGCNA analysis across anatomical regions.
**Figure S8**. Comparative analysis of T21 RNA-seq and microarray datasets.
**Figure S9**. Loess fold-change plots across the genome.
**Figure S10**. Percentage of differentially expressed genes by chromosome.
**Figure S11**. Transcriptomic concordance across multiple trisomy 21 RNA-seq investigations.
**Figure S12**. Correlation between gene expression and RNA editing levels.
**Figure S13**. COG3 and RNA recoding percent change values.
**Figure S14**. Editing concordance with cell type-specific and fresh brain RNA-seq investigations.
**Figure S15.** RNA editing in Trisomy 21 immune cells.

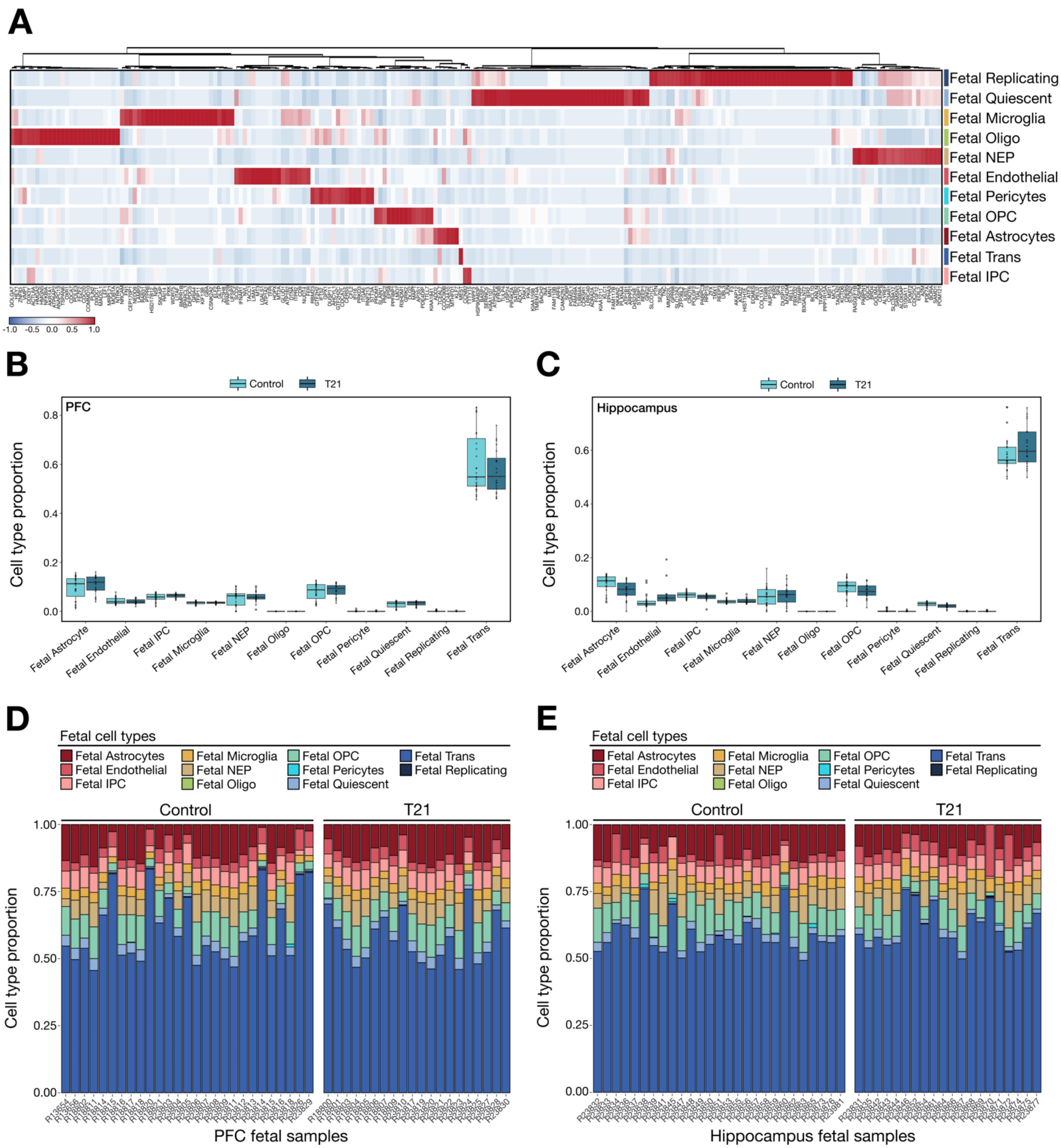


**Figure S1.** Cell type deconvolution of fetal brain RNA-seq data. (**A**) Heatmap displaying the expression of marker genes used in the reference signature matrix from Pei et al., 2021 across 11 mid-gestational human brain cell types. Marker expression is z-score normalized and hierarchically clustered to confirm specificity across cell populations, including fetal astrocytes, endothelial cells, microglia, NEP (neuroepithelium), OPC (oligodendrocyte precursor cells), pericytes, oligodendrocytes, IPC (intermediate progenitor cells), quiescent, replicating, and transitional neurons. (**B-E**) Cell-type fractions were estimated using the est_frac function from bMIND (v1.2.0), which uses non-negative least squares within a Bayesian framework to deconvolve bulk RNA-seq data, guided by prior cell-type signatures from Pei et al. (2021). (**B–C**) Boxplots showing estimated cell type proportions in PFC (B) and hippocampus (**C**) for T21 and euploid control samples. Cell fractions were computed via non-negative least squares regression using the MIND algorithm, applied to $\log_2$-transformed CPM values. Each box represents the distribution of estimated proportions across samples for a given cell type and diagnosis group. (**D–E**) Stacked barplots visualizing estimated cell type composition per sample in PFC (D) and hippocampus (E), stratified by diagnosis (Control vs. T21). Each bar represents the sum-normalized proportions of the 11 cell types in an individual sample. T21 and control samples exhibit comparable cellular composition, with minimal evidence of major compositional shifts.

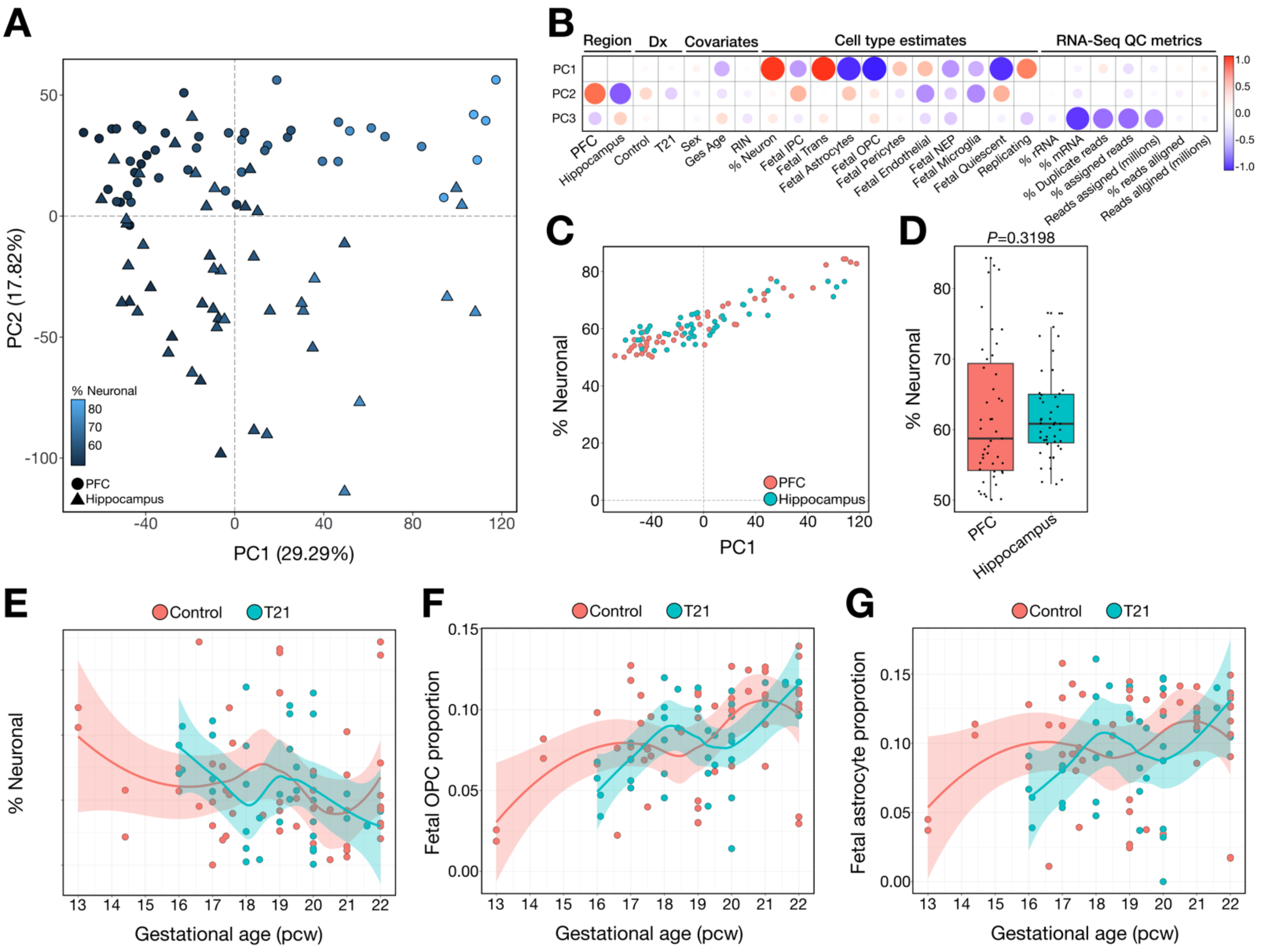


**Figure S2**. **Quality control of normalized fetal brain RNA-seq data.** (**A**) Raw gene-level expression counts were first filtered to exclude lowly expressed genes and then normalized using the voom transformation from the limma package, which models the mean–variance relationship in log-transformed count data and generates precision weights for linear modeling. These voom-normalized values were used for Principal Component Analysis (PCA), visualized separately for dorsolateral prefrontal cortex (PFC, circles) and hippocampus (triangles) samples, with color indicating estimated neuronal fraction. PC1 and PC2 account for 29.3% and 17.8% of the total variance, respectively, and reveal clear regional separation, with neuronal content as a major driver of transcriptomic variation. (**B**) Correlation heatmap showing associations between the top three principal components and metadata variables, including brain region, diagnosis (Dx), biological covariates (e.g., age, sex), estimated cell type proportions, and RNA-seq quality control (QC) metrics. Circle size and color indicate strength and direction of Pearson correlations (blue = negative; red = positive). PC1 strongly correlates with brain region and neuronal proportion. (**C**) Scatterplot showing % neuronal composition versus PC1, stratified by brain region. Neuronal proportion closely tracks with PC1 values, highlighting its influence on transcriptomic variation. (**D**) Boxplot comparing estimated % neuronal content between PFC and hippocampus samples. While mean neuronal proportions appear lower in hippocampus, differences are not statistically significant (Wilcoxon P = 0.3198). (**E**–**G**) Scatterplots showing relationships between gestational age and estimated cell type proportions for neuronal cells (**E**), oligodendrocyte precursor cells (OPCs, **F**), and astrocytes (**G**), stratified by diagnosis (Control vs. T21).

(**E-F**) LOESS regression lines with 95% confidence intervals were used for visualizing cell type proportions over developmental trajectories because they do not assume a predefined parametric form and instead flexibly fit local subsets of the data; especially valuable when cell type dynamics exhibit inflection points or vary across tissue types and conditions. Developmental trajectories in cell composition are broadly similar across groups, though subtle deviations in T21 samples are observed in select cell types.

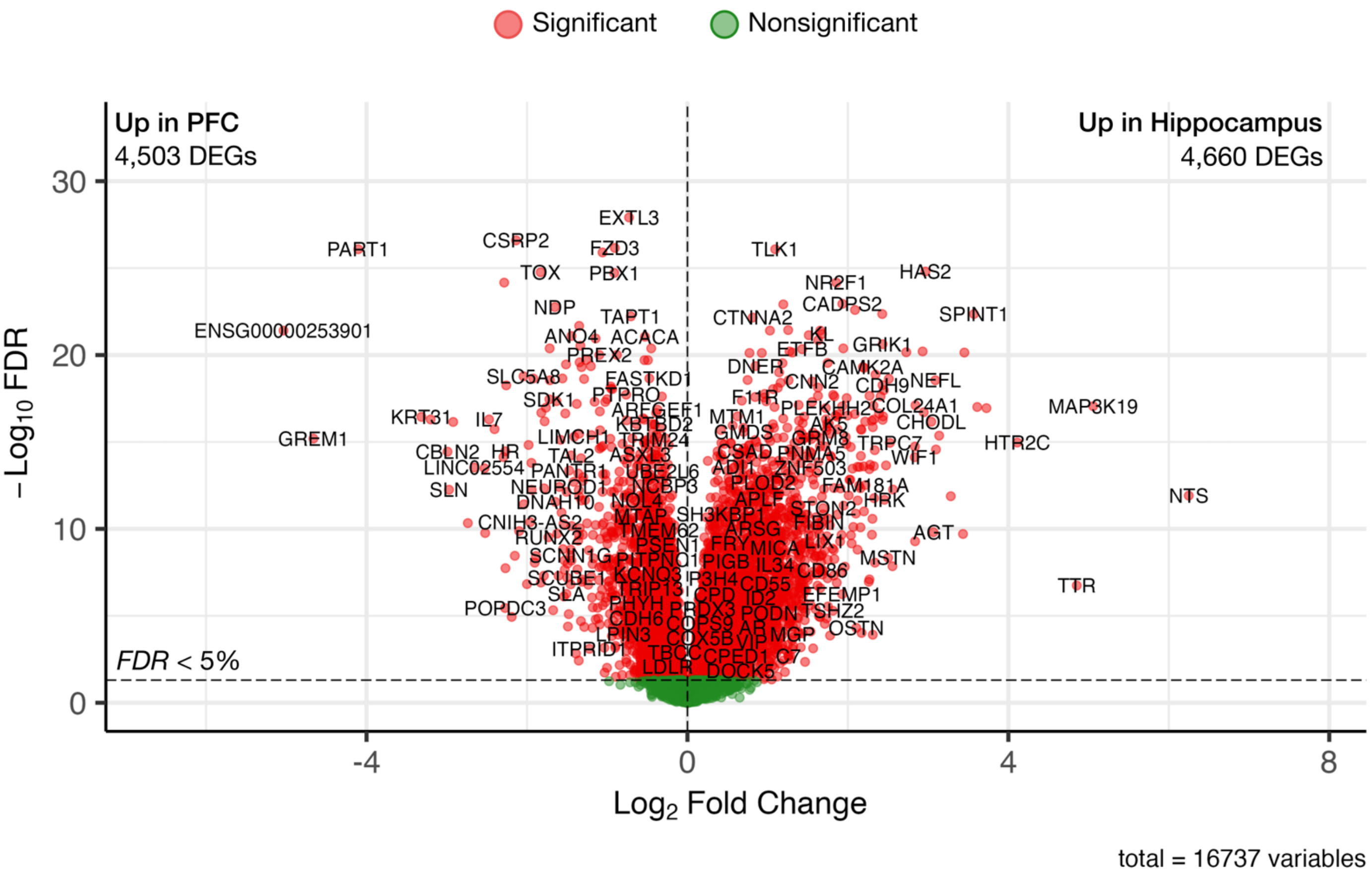


**Figure S3**. **Differential gene expression between anatomical regions**. Volcano plot illustrating transcriptome-wide differential gene expression between prefrontal cortex (PFC) and hippocampus in mid-gestational human fetal brain ($n = 47$ individuals; paired samples). Each point represents a gene, with the x-axis denoting $\log_2$ fold change (positive = upregulated in hippocampus, negative = upregulated in PFC) and the y-axis indicating statistical significance ($-\log_{10}$ FDR-adjusted p-value). Genes meeting a false discovery rate (FDR) threshold of <5% are shown in red (Significant), while non-significant genes are in green. A total of 9,163 genes were differentially expressed between regions (4,503 upregulated in PFC; 4,660 upregulated in hippocampus). Key marker genes with strong region-specific expression are labeled. The dashed horizontal line marks the FDR = 0.05 threshold.

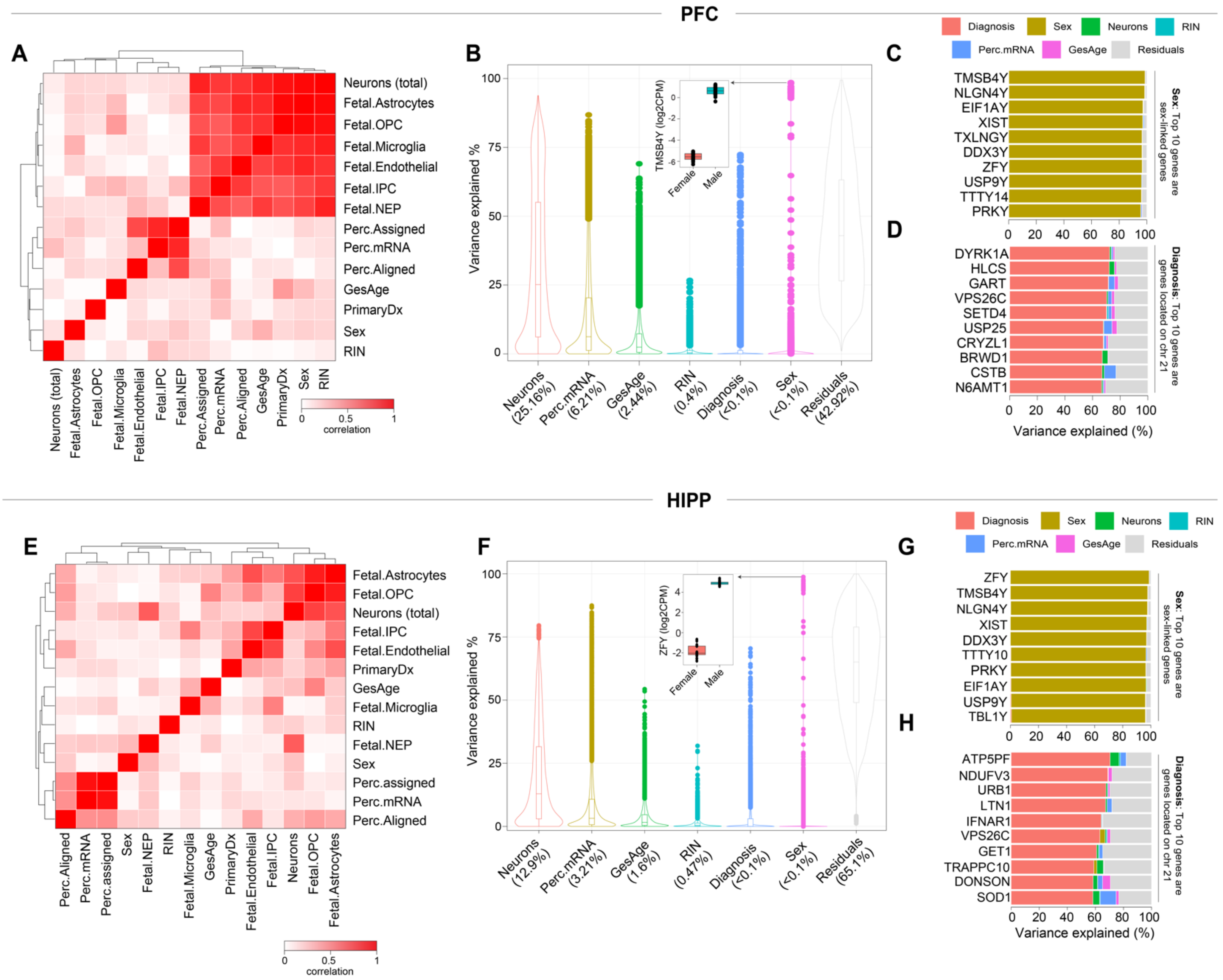


**Figure S4. Variance of gene expression explained within each region.** Correlation heatmaps displaying relationships among technical, biological, and cellular covariates in PFC (**A**) and hippocampus (**E**). Variables include neuronal proportion, cell-type fractions (e.g., fetal astrocytes, OPCs), RNA integrity number (RIN), gestational age (GesAge), percent mRNA, and sex. (**B, F**) Violin plots depicting the proportion of gene expression variance explained by each covariate in the PFC (**B**) and hippocampus (**F**), based on linear mixed models (variancePartition). Individual points represent single genes, with horizontal lines denoting median values. (**C, G**) Top 10 genes in each region whose expression is most strongly associated with sex. (**D, H**) Top 10 genes whose expression is most strongly associated with T21 diagnosis. Bars are color-coded by the proportion of variance explained by each factor. Overall, neuronal composition and percent mRNA were the dominant contributors to expression variance, while diagnosis accounted for substantial variance in select T21-sensitive genes.

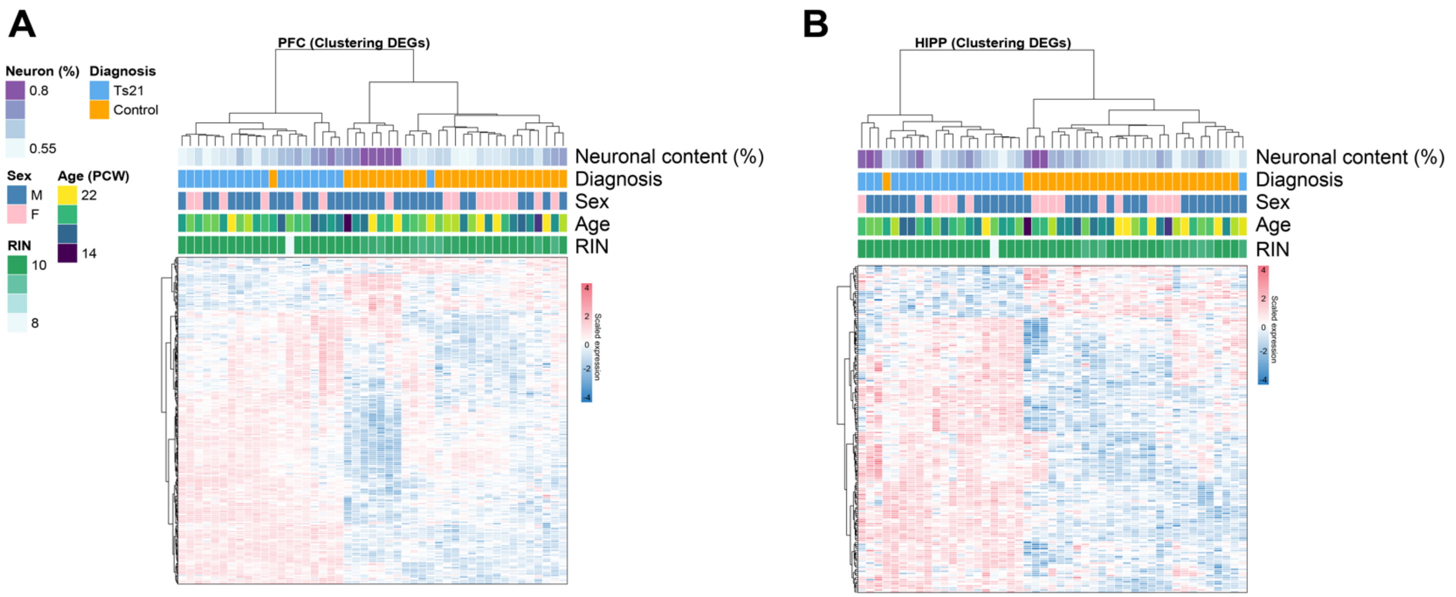


**Figure S5**. **Unsupervised clustering of differential gene expression signatures.** Heatmaps displaying unsupervised hierarchical clustering of differentially expressed genes (DEGs) in the dorsolateral prefrontal cortex (PFC; **A**) and hippocampus (HIPP; **B**). Gene expression values were scaled and centered prior to clustering. Columns represent individual samples, annotated by neuronal content (%), T21 diagnosis (purple = T21; orange = control), sex (blue = male; pink = female), gestational age in post-conception weeks (PCW), and RNA integrity number (RIN). Rows represent DEGs identified at FDR < 5% within each region. Clustering reveals distinct expression profiles that align with diagnosis and neuronal proportion, suggesting robust T21-associated transcriptional changes.

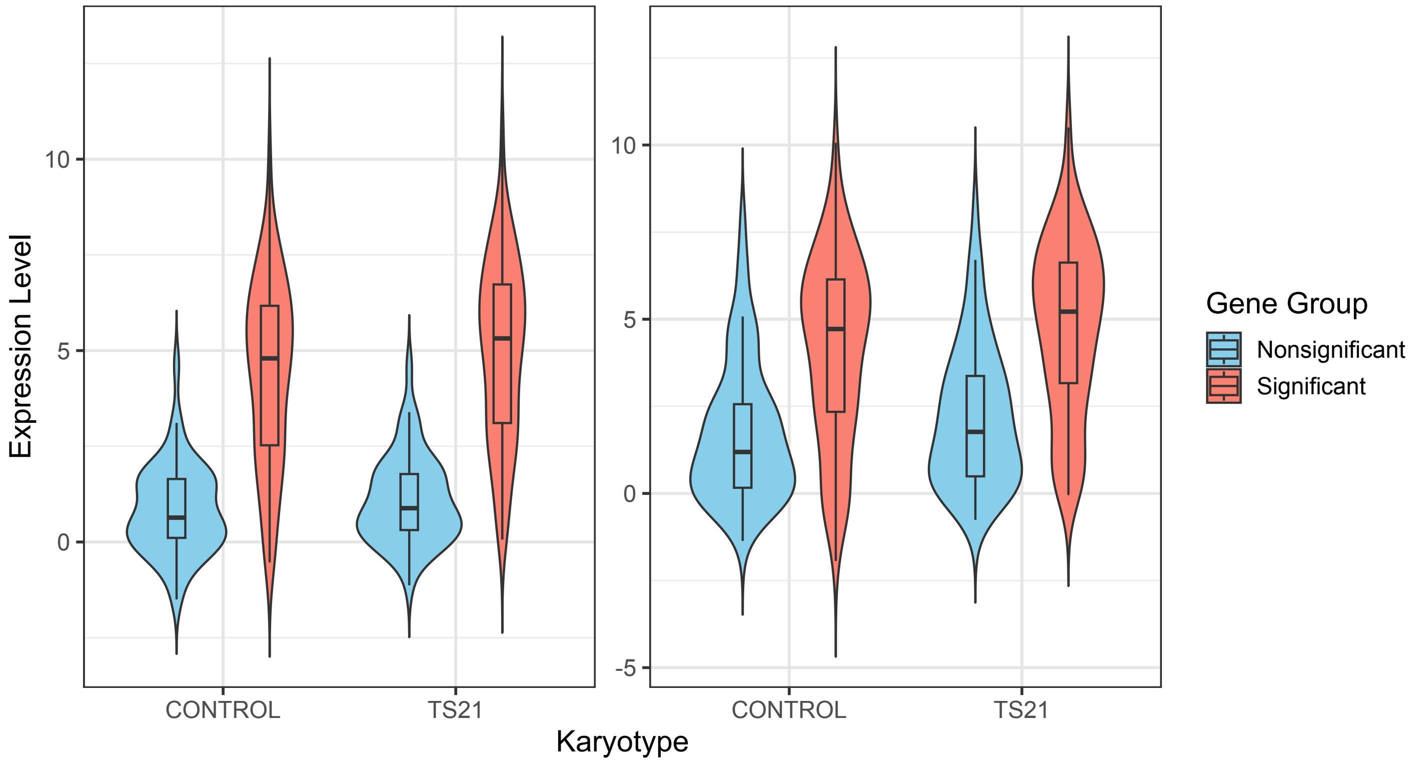


**Figure S6. Expression levels of dosage-sensitive versus non-sensitive chromosome 21 genes across control and T21 brain samples.** Violin and box plots show normalized gene expression levels ($log_2$ CPM) for chromosome 21 genes that were significantly upregulated in T21 cases ("Significant") versus those without differential expression ("Nonsignificant"). Data are shown separately for control and T21 samples, stratified by brain region (PFC, left; hippocampus, right). In both regions, significantly upregulated genes in T21 were more highly expressed at baseline in control samples (PFC: FC = 2.29, $p = 1.1\times10^{-10}$; hippocampus: FC = 2.31, $p = 1.3\times10^{-11}$; Wilcoxon rank-sum test), suggesting that genes with higher basal expression are more likely to exhibit dosage sensitivity in the trisomic brain.

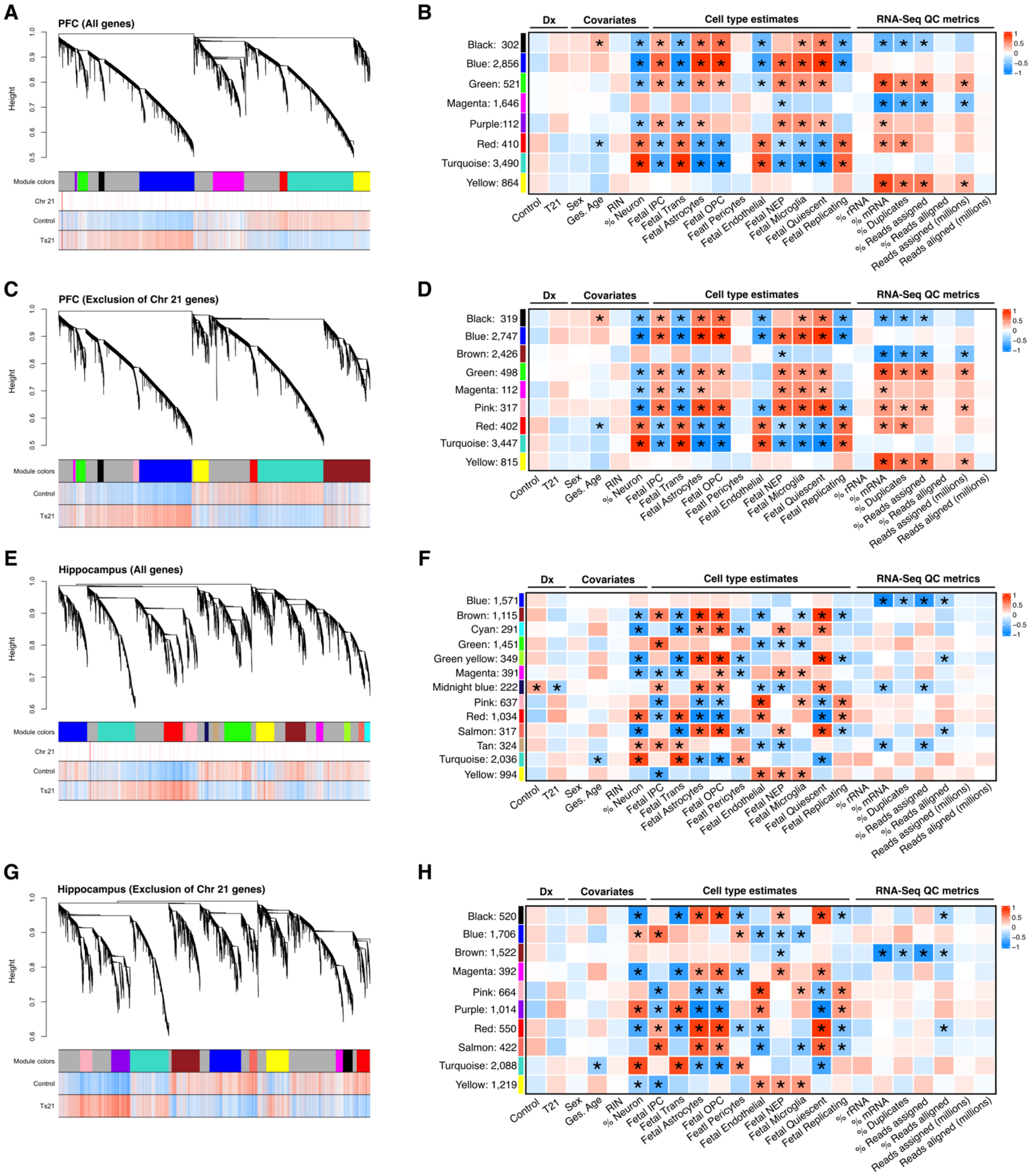


**Figure S7. Co-expression network structure and module–trait associations across brain regions.** (**A–B**) WGCNA in PFC using all expressed genes. (**A**) Gene dendrogram with module assignments; annotation bars indicate chromosome 21 membership, T21 status, and controls. (**B**) Module eigengene correlations with diagnosis, cell type estimates, and RNA-seq quality metrics (FDR-adjusted $p < 0.05$ indicated by asterisks). (**C–D**) PFC WGCNA excluding chromosome 21 genes, showing revised module structure (**C**) and trait associations

(**D**). (**E–F**) WGCNA in hippocampus using all genes, with corresponding dendrogram (**E**) and trait associations (**F**). (**G–H**) Hippocampus analysis excluding chromosome 21 genes, with updated module structure (**G**) and eigengene correlations (**H**). These analyses evaluate the contribution of chromosome 21 genes to network architecture and identify region-specific co-expression modules associated with T21 and key covariates.

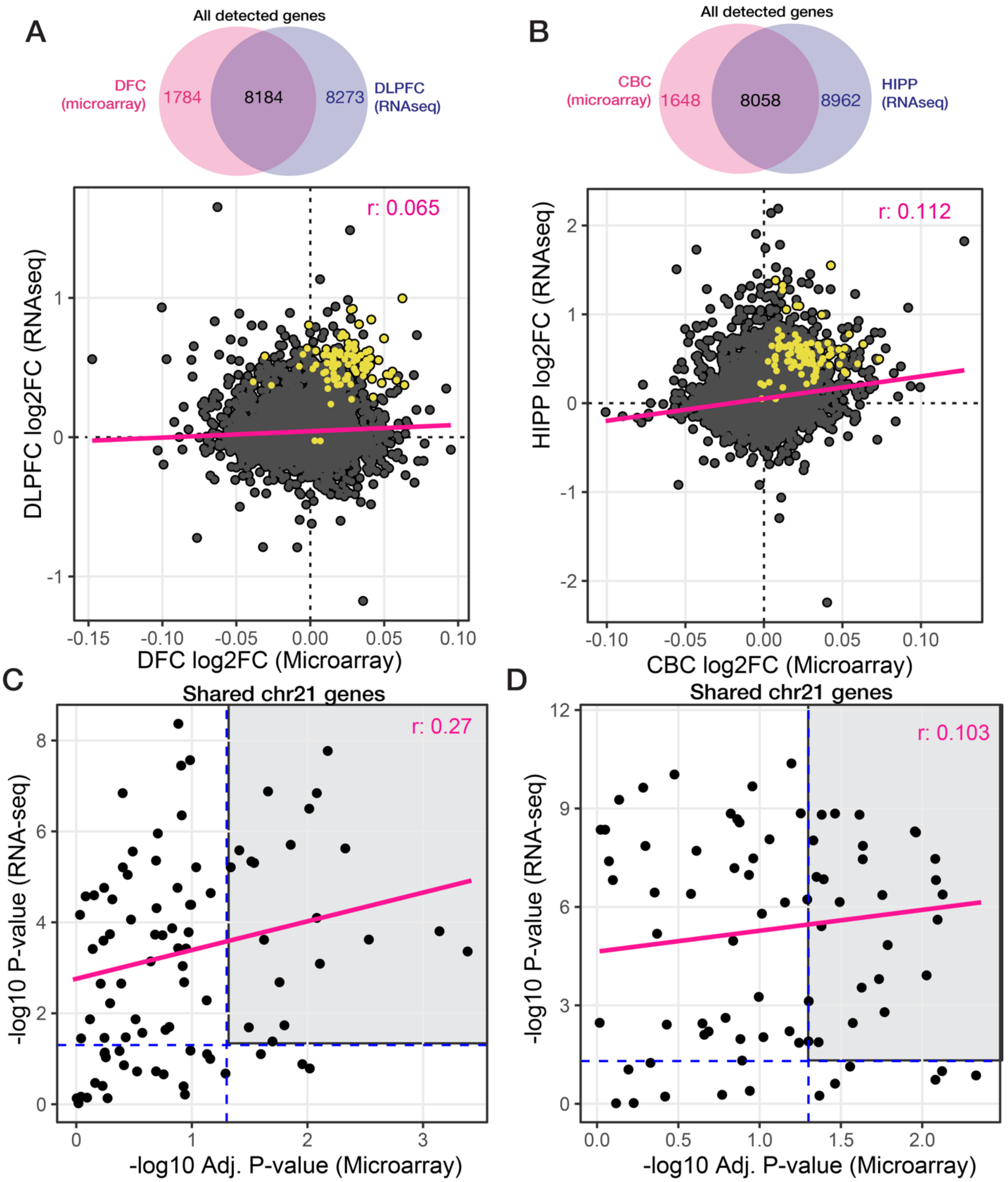


**Figure S8**. **Comparative analysis of T21 RNA-seq and microarray datasets.** Comparing our fetal brain RNA-seq data to matched regions from a previously published microarray study (Olmos-Serrano et al., 2016). (**A**) Comparison between detected genes (**A**) in the dorsolateral prefrontal cortex (PFC, RNA-seq) and DFC (microarray) and (**B**) the hippocampus (HIPP, RNA-seq) and CBC (microarray). Top Venn diagrams depict overlap in detected genes between platforms. Bottom scatterplots depict the log2 fold-changes in the microarray (y-axes) relative to RNA-seq data (x-axes). Pink lines represent the linear regression fit; Pearson correlation coefficients (r) are shown in pink. (**C–D**) scatterplots depict the log2 fold-changes for chromosome 21 genes in the microarray (y-axes) relative to RNA-seq data (x-axes) – (**C**) PFC vs. DFC; (**D**) HIPP vs. CBC. Blue dashed lines indicate significance thresholds (FDR < 5%). Correlation values highlight modest, but positive, concordance for chromosome 21 genes, especially in the PFC. These analyses support cross-platform validation of key dosage-sensitive transcripts in trisomy 21.

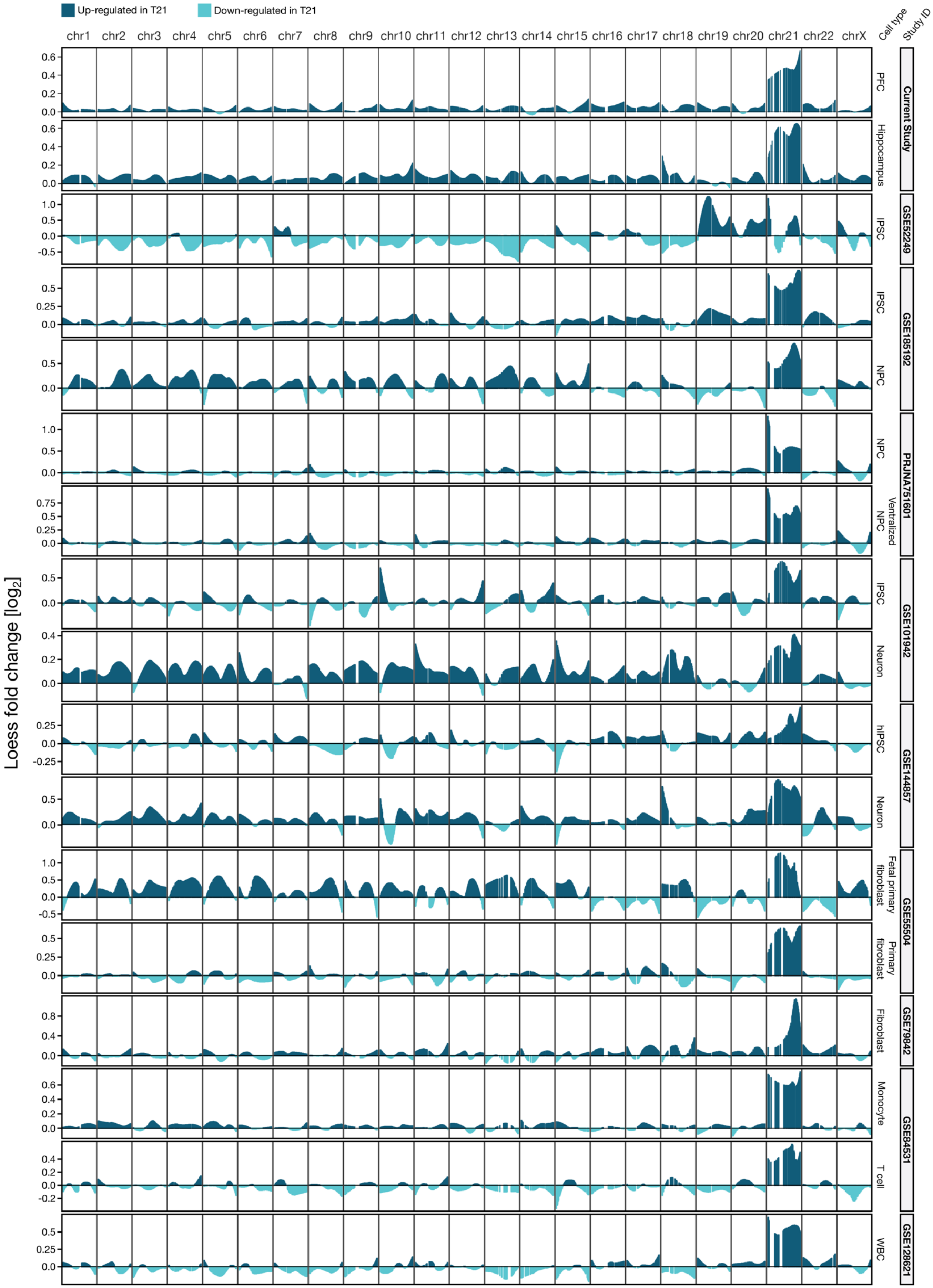


Loess $log_2$[FC] values are plotted based on the gene positions along the chromosomes.

**Figure S9**. **Loess fold-change plots across the genome.** Loess-smoothed $\log_2$ fold-change values are plotted for each chromosome across 17 datasets comparing trisomy 21 (T21) to euploid control samples. Each row represents a unique dataset or cell type, including fetal PFC and hippocampus from this study (top two rows), as well as independent RNA-seq datasets from diverse cell types (iPSCs, NPCs, neurons, fibroblasts, T cells) and tissues spanning developmental time points (listed at right; see Table 1 for full metadata). For each gene, the genomic coordinate was used to order $\log_2$ fold-change values along the x-axis, and Loess smoothing was applied to visualize local patterns of up- (blue) and down-regulation (aqua) across the genome. Robust and consistent overexpression is observed across the chromosome 21 (chr21) region in nearly all datasets, consistent with 1.5× gene dosage effects in T21. Localized downregulation is also observed on non-chr21 chromosomes, suggesting potential compensatory or secondary transcriptional responses.

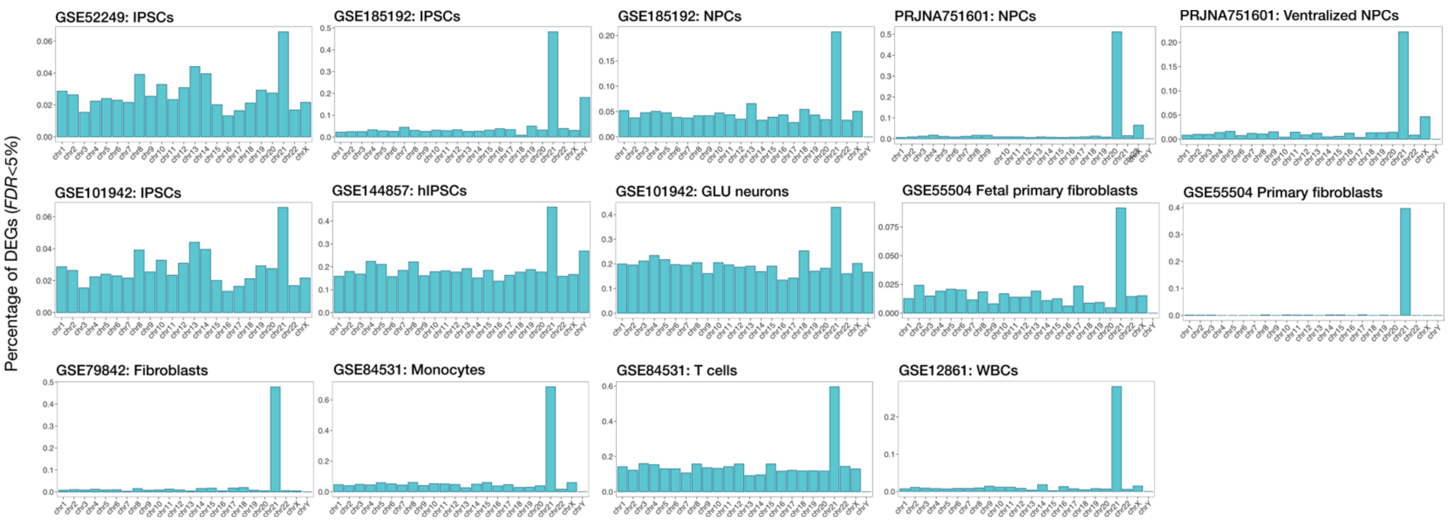


**Figure S10. Percentage of differentially expressed genes by chromosome across datasets.** Bar plots show the percentage of genes per chromosome that were differentially expressed (FDR < 5%) in trisomy 21 (T21) versus control comparisons across 15 independent RNA-seq datasets, including iPSCs, neural progenitor cells (NPCs), glutamatergic neurons, fibroblasts, monocytes, T cells, and white blood cells (WBCs). Each panel corresponds to a unique dataset and cell type (see Table 1 for details). The y-axis indicates the proportion of DEGs relative to the total number of detected genes on each chromosome. A strong and consistent enrichment of differentially expressed genes on chromosome 21 is observed across most datasets, consistent with primary dosage effects in T21. Varying levels of differential expression across other chromosomes likely reflect cell type–specific transcriptional responses or technical variation.

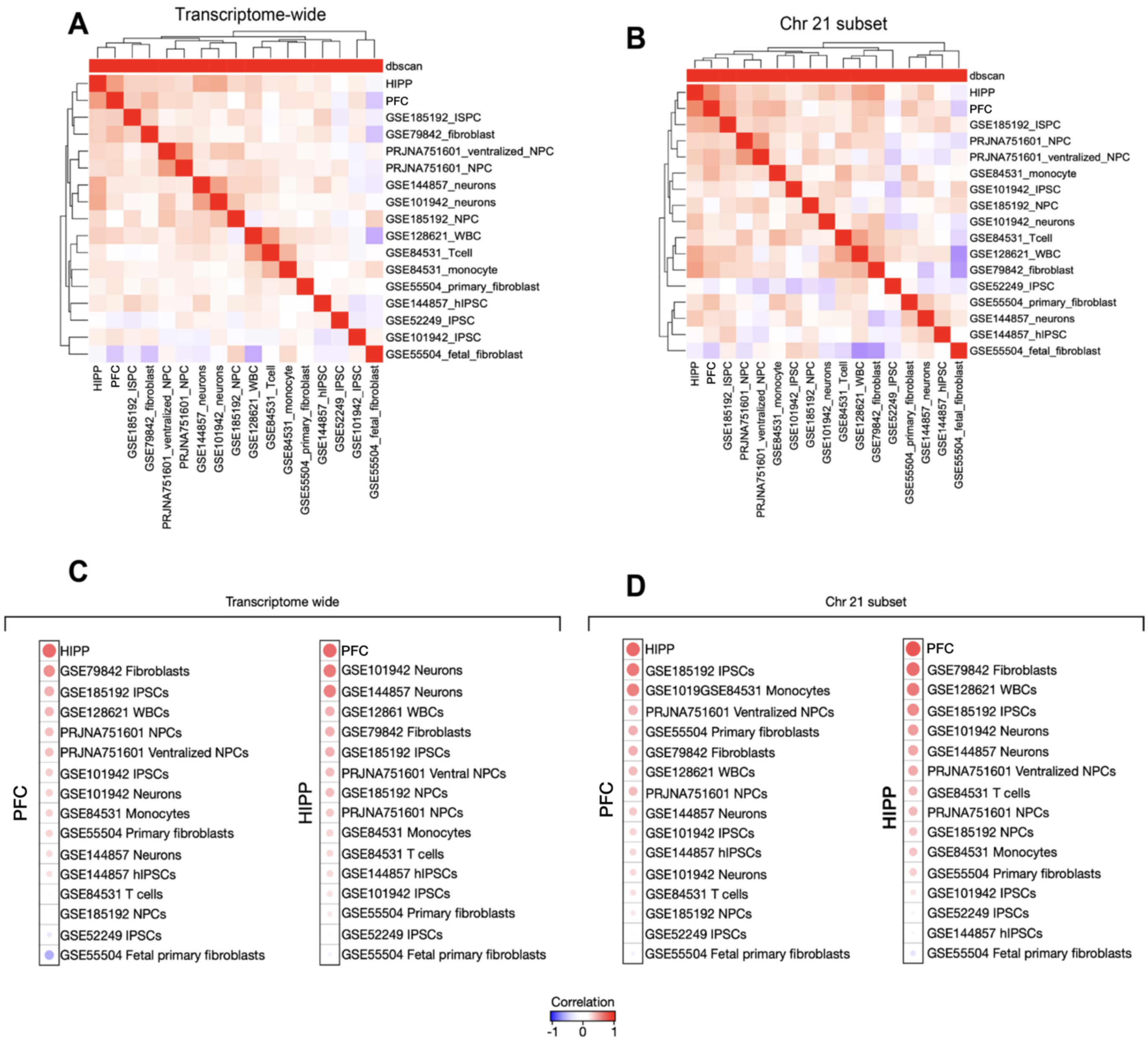


**Figure S11. Transcriptomic concordance across multiple T21 RNA-seq investigations.** (**A–B**) Pairwise Pearson correlation heatmaps of $log_2$ fold-change values comparing PFC and hippocampus DEGs from this study to 15 external RNA-seq datasets of T21 versus control. (**A**) Transcriptome-wide analysis across all detected genes. (**B**) Analysis restricted to chromosome 21 genes. Red indicates strong positive correlation in differential expression signatures. Clustering reveals closer concordance among neural and progenitor cell datasets, particularly with fetal cortex. (**C–D**) Ranked correlation dot plots of external datasets with PFC and hippocampus DEGs. (**C**) Full transcriptome correlations. (**D**) Chromosome 21–restricted correlations. Dot size reflects correlation magnitude; all points are color-coded by correlation direction. Notably, datasets derived from cortical neurons, iPSCs, NPCs, and fetal fibroblasts show moderate concordance with primary fetal brain tissue. These findings highlight reproducible gene expression signatures in T21 across diverse models, with strongest consistency observed on chromosome 21.

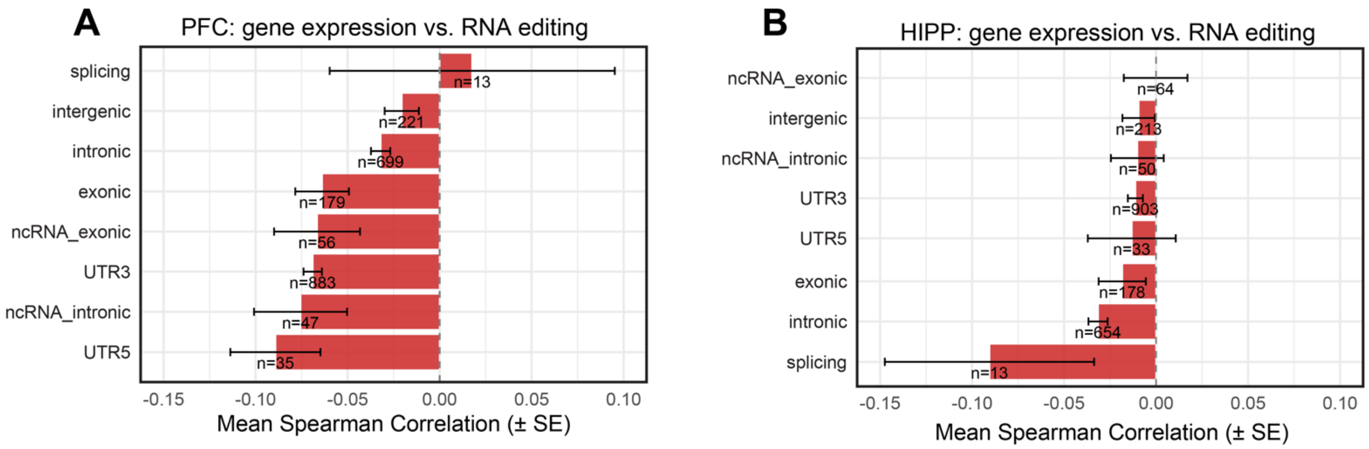


**Figure S12. Inverse coupling between RNA editing and gene expression across genic regions in human PFC and hippocampus (HIPP).** Bar plot showing the mean Spearman correlation coefficients (± standard error) between RNA editing levels and gene expression across all genes, stratified by genic region of the editing sites (e.g., 3′ UTR, 5′ UTR, intronic, exonic, intergenic). Each correlation was computed across matched subjects for a given gene and its corresponding editing sites, using bulk RNA-seq from (**A**) prefrontal cortex (PFC) and (**B**) hippocampus. All correlations are negative, consistent with an inverse relationship between A-to-I editing and transcript abundance. Numbers next to each bar indicate the number of gene–region pairs contributing to each summary. The dashed line denotes zero correlation for reference.

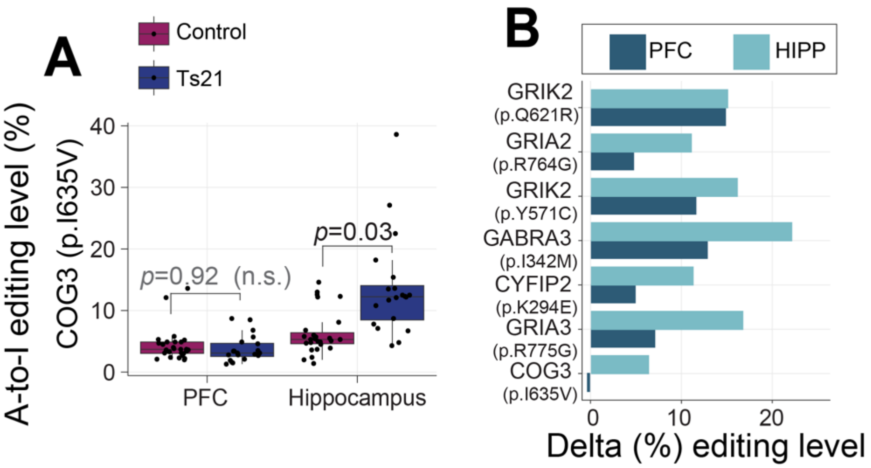


**Figure S13. COG3 editing differences and RNA recoding dynamics in T21**. (**A**) A-to-I RNA editing levels at the canonical recoding site COG3 (p.I635V) in control and T21 fetal brain samples. Boxplots show editing levels stratified by diagnosis and brain region. A significant increase in editing is observed in the hippocampus ($p = 0.03$), but not in the PFC ($p = 0.92$; not significant, n.s.). (**B**) Barplot showing the difference in percent editing levels (Δ editing) for eight canonical A-to-I recoding sites between T21 and control samples across PFC and hippocampus. Genes include subunits of glutamate receptors (GRIA2, GRIA3, GRIK2), a GABA receptor (GABRA3), a vesicle trafficking factor (COG3), and other functionally conserved recoding targets. Values represent average editing level differences by region.

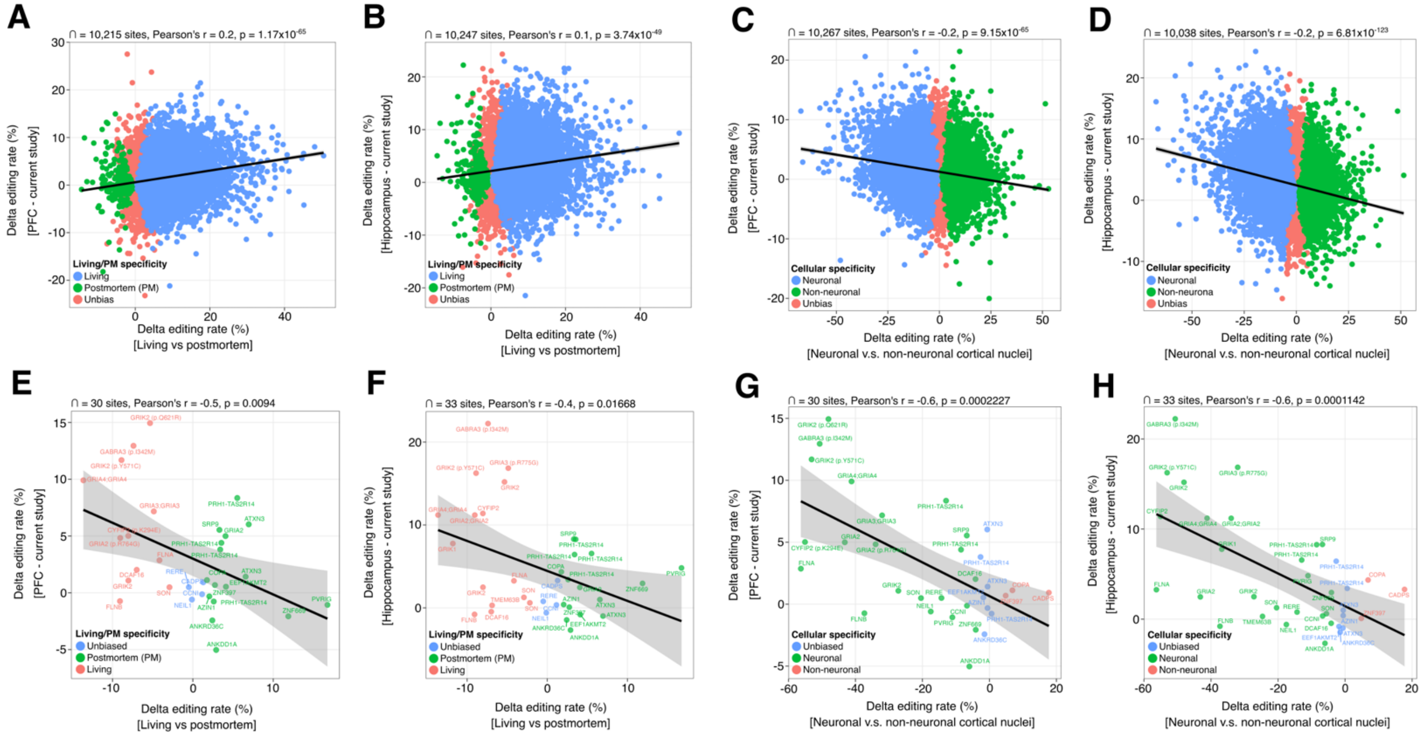


**Figure S14. Editing concordance with cell type-specific and fresh brain RNA-seq investigations.** (**A–B**) Scatterplots comparing delta editing rates (T21 vs. control) in PFC (**A**) and hippocampus (**B**) from the current study to editing differences between living vs. postmortem (PM) human brain samples, stratified by site-level specificity (Living, PM, or Unbiased). Positive correlations suggest partial alignment of editing changes with RNA integrity and PMI effects. (**C–D**) Same as (**A–B**), but plotted against editing changes between neuronal and non-neuronal nuclei, highlighting cellular specificity effects. (**E–F**) Subset analysis of 30–33 curated recoding sites for PFC (**E**) and hippocampus (**F**), showing negative correlations between current study delta editing and postmortem-associated editing. (**G–H**) Subset analysis for the same sites, comparing delta editing in our dataset with neuronal vs. non-neuronal nuclei differences. Strong negative correlations suggest that sites normally enriched in neurons are hypoedited in T21, consistent with reduced neuronal proportion. Linear regression lines with 95% confidence intervals are shown.

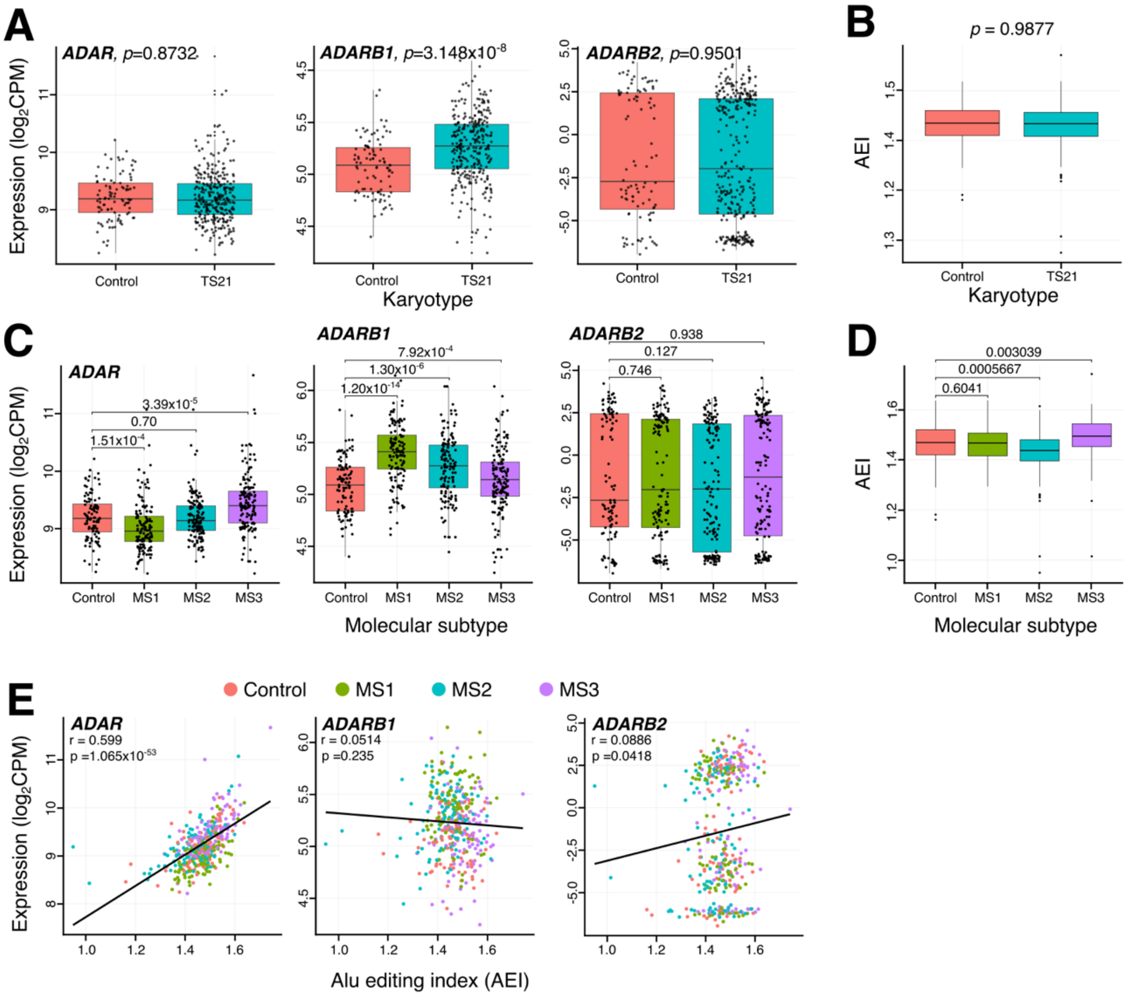


**Figure S15. RNA editing and ADAR expression in Trisomy 21 immune cells by molecular subtype.** (**A**) Expression levels (log2CPM) of ADAR, ADARB1, and ADARB2 in euploid controls and individuals with Trisomy 21 (TS21), compared using Wilcoxon rank-sum tests. ADARB1 expression is significantly elevated in TS21. (**B**) *Alu* Editing Index (AEI), a global measure of A-to-I RNA editing, shows no significant difference between TS21 and controls. (**C**) Expression levels of ADAR family genes across TS21 molecular subtypes (MS1–MS3) and controls. ADAR and ADARB1 levels are significantly elevated in MS2 and MS3. (**D**) AEI stratified by molecular subtype reveals increased global RNA editing in MS2 and MS3 compared to controls. **(E)** Correlation between AEI and gene expression for ADAR (left), ADARB1 (middle), and ADARB2 (right). Only ADAR expression is strongly positively correlated with AEI ($r = 0.599$, $p = 1.06 \times 10^{-30}$), consistent with its role as the primary mediator of global editing levels in immune cells.
